# Pupil Abnormality Frequency in the First 72 Hours Improves IMPACT score in Traumatic Brain Injury

**DOI:** 10.1101/2024.11.23.24317826

**Authors:** Divya Veerapaneni, Naveen Arunachalam Sakthiyendran, So Yeon Kim, Chuong Nguyen, Ali Daneshmand, Mohamad Abdalkader, Shariq Mohammed, Josée Dupuis, Kevin N. Sheth, Emily J. Gilmore, David Greer, Charlene J. Ong

**Author notes:** **Corresponding author:** Charlene J. Ong, MD, MPHS Assistant Professor of Neurology Boston University Chobanian and Avedisian School of Medicine 85 East Concord St, Boston, MA 02118. **Conflict of interest disclosure:** The authors declare no conflicts of interest. This report was prepared according to Strengthening the Reporting of Observational Studies in Epidemiology reporting guidelines. C.J.O. receives support from National Institutes of Health (NIH)/National Institute of Neurologic Disorders and Stroke K23NS116033 for this study. This manuscript has not been published and is not under consideration elsewhere. This study is adherent to ethical guidelines and approved by the Boston Medical Center Institutional Review Board. Informed consent was waived due to the anonymous nature of the study.

## Abstract

**Importance:** In patients with traumatic brain injury (TBI), baseline pupillary assessment is common. However, the incidence and frequency of pupil abnormalities within the first several days remain poorly characterized.

**Objectives:** Our aim was to test the association between pupil abnormality frequency over the first 72 hours of admission and clinical outcomes.

**Design:** We conducted a retrospective observational study of patients with a primary diagnosis of TBI with at least three quantitative pupillometry measurements within 72 hours at a single-center ICU from 2018 to 2022.

**Outcomes and Measures:** Neurological Pupil index (NPi), a quantitative composite metric for pupil reactivity, was obtained at each clinical neurologic assessment over 72 hours. Pupil measurements were defined as abnormal if they had a NPi of <3 in either eye, NPi asymmetry ≥0.7, or pupil size asymmetry ≥1mm. We tested the association of increased frequency of pupil abnormalities over 72 hours and unfavorable discharge disposition (death, hospice, or long-term care) using multivariable logistic regression, adjusting for confounders. We then compared whether the IMPACT model was improved by the frequency of pupil abnormalities using goodness-of-fit.

**Results:** Of 131 patients, median age was 59 years, and 30% were women. Thirty-five percent had unfavorable discharge disposition. Pupil abnormalities occurred in 62%, 61%, and 88% of mild, moderate, and severe TBI patients, respectively. Odds ratio of unfavorable discharge for every 1% increase in pupil abnormality frequency was 1.03 (95% Cl, 1.01-1.05), equivalent to one additional abnormal pupil measurement within a 72-hour period. The adjusted IMPACT TBI model’s goodness-of-fit improved with pupil abnormality frequency (*X*^2^=5.67, *p*=0.02).

**Conclusions and Relevance:** Pupil abnormalities occur commonly in TBI and have the highest frequency in severe TBI. Increased pupil abnormality frequency is associated with unfavorable discharge disposition and improves performance of prognostic TBI models.

**Key Points:** The goal of this study was to test the association between pupil abnormality frequency over the first 72 hours of admission and unfavorable discharge disposition and whether it improved the IMPACT model. In this retrospective observational study of 131 TBI patients at a single-center, we found that an increased frequency of pupil abnormalities across 72-hours significantly correlated with unfavorable discharge in patients and improved the IMPACT model’s goodness-of-fit. These findings highlight the potential of long-term pupillary metrics and their role as not only a prognostic indicator in patients but also a tool that improves the performance of prognostic TBI models.

## Background

Traumatic brain injury (TBI) is a leading cause of mortality and morbidity worldwide.^1^ Absent unilateral or bilateral pupil reactivity on admission is associated with worse outcomes in TBI patients^2,3^ and is used in the IMPACT TBI model,^4,5^ a well-established model for predicting clinical outcomes in patients with moderate and severe TBI. Moreover, close neurologic monitoring including subjective pupil size and reactivity assessment is used to inform clinical decision-making for TBI patients during admission.^6,7^ However, quantification of pupil abnormalities over time and between TBI severity groups is not well characterized despite the increasing availability of quantitative pupillometers in intensive care settings.^9^ Moreover, while prior studies have demonstrated an association between reduced pupil reactivity and long term outcome in heterogenous acute brain injury patients including TBI,^10^ it is unclear whether inclusion of longitudinal pupillometry can improve on existing prediction scales. Understanding the expected frequency of pupil abnormalities and their association with clinical outcomes in the first 72 hours post-TBI may improve detection of early neurologic deterioration and outcome prognostication.

Our aim was to determine the prognostic value of pupil abnormality frequency by 1) characterizing the prevalence of pupil abnormalities within different TBI severities 2) testing its association with clinical outcomes and 3) investigating whether pupil abnormality frequency improves existing TBI prognostic models. We hypothesized that increased pupil abnormality frequency is associated with worse outcomes and that the inclusion of pupil abnormality frequency over the first 72 hours of admission improves TBI prognostication.

## Methods

### Study Population

We conducted a single-center retrospective study of TBI patients admitted to the Boston Medical Center surgical and neurological intensive care units between 2018 and 2022 and who had at least three quantitative pupil measurements collected within the first 72 hours of hospitalization. Exclusion criteria included pre-existing eye conditions that may impact pupil reactivity such as diabetic retinopathy and glaucoma (**eFigure 1**).

**Figure 1:**
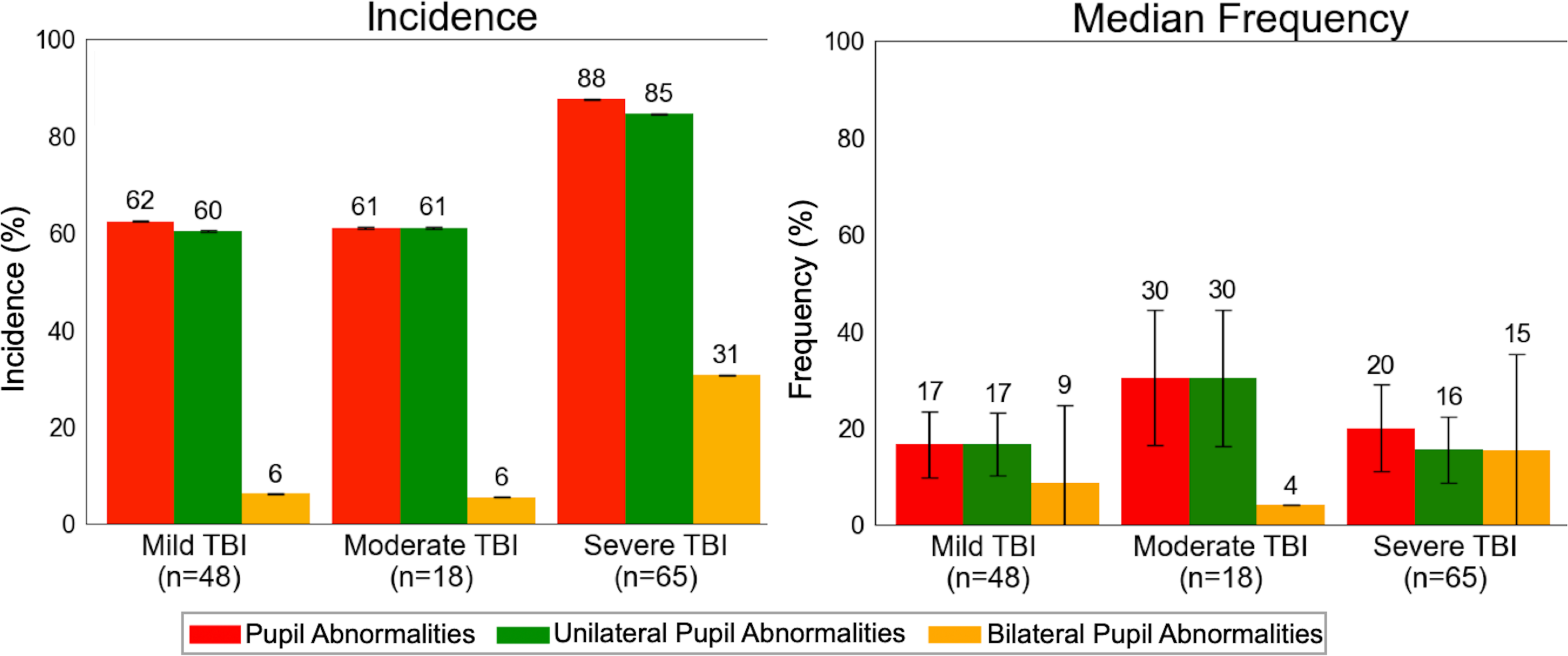
Percent Incidence and Median Frequency of Pupil Abnormalities between Traumatic Brain Injury Severity Groups (*n* = 131) Percent incidence and median frequency of pupil abnormalities. Incidence and median frequency of unilateral pupil abnormalities were higher compared to bilateral pupil abnormalities across most TBI severity groups. Frequency was reported as percentage and includes only patients with nonzero frequencies. **Abbreviations:** TBI = Traumatic Brain Injury, % = Percent **Pupil Abnormalities:** Neurological Pupil index (NPi) < 3, NPi difference between both eyes (Diff NPi) ≥ 0.7, or Size difference between both eyes (Diff Size) ≥ 1mm **Unilateral Pupil Abnormalities:** Unilateral NPi < 3, Diff NPi ≥ 0.7, or Diff Size ≥ 1mm **Bilateral Pupil Abnormalities:** Bilateral NPi < 3

### Data Collection

We collected demographic and clinical information from the electronic health record through the Clinical Data Warehouse (**eMethods**). Using the admission Glasgow Coma Scale (GCS) in the intensive care unit post-resuscitation, patients were categorized as mild (GCS 13-15), moderate (GCS 9-12), or severe (GCS < 9) TBI.^11^ For each patient, we measured radiographic features including midline shift and effacement of basal cisterns from the most severe brain CT in the first 72 hours to compute the Marshall CT score,^12^ a radiographic classification system for TBI, after establishing interrater reliability of at least 0.70 (95% CI, 0.42-0.99). The most severe brain CT in the first 72 hours was used for the analysis to reflect the overall severity of the patient’s condition after potential deterioration rather than the patient’s baseline at admission. Trained nursing staff collected pupillometry data using the NeurOptics NPI-300^TM^ pupillometer (NeurOptics Inc., Irvine CA, USA) during standard neurological checks (every one, two, or four hours). The pupillometer reports the Neurological Pupil index (NPi), a composite score of pupil reactivity based on resting and constricted pupil size, percent change, constriction velocity, dilation velocity, and latency. Scores range from 0 to 5, with values < 3 considered abnormal by the manufacturer. This report was prepared according to Strengthening the Reporting of Observational Studies in Epidemiology reporting guidelines.^13^

### Outcomes

The primary outcome was unfavorable discharge disposition, defined as death, hospice, or long-term facility placement. Other outcomes included death at discharge, and documented evidence of eye tracking at discharge (**eMethods**). Outcomes were determined from discharge summaries and ascertained by two reviewers (C.J.O. and D.V).

### Exposures

The primary exposure was pupil abnormality frequency calculated per patient and defined as the percentage of abnormal pupil measurements over 72 hours. Pupils were considered “abnormal” if NPi < 3 in either eye, NPi asymmetry between eyes (Diff NPi) ≥ 0.7, or resting pupil size asymmetry between eyes (Diff Size) ≥ 1mm. Definitions were based on manufacturer recommendations (NPi < 3 and Diff NPi ≥ 0.7) and prior work on anisocoria in the neuro-ICU (size ≥ 1mm).^14^ Other metrics including constriction velocity, dilation velocity, and latency are encompassed in the NPi variable. Secondary exposures were frequencies of unilateral and bilateral pupil abnormalities. Unilateral pupil abnormalities were defined as unilateral NPi < 3, Diff NPi ≥ 0.7, or Diff Size ≥ 1mm, while bilateral pupil abnormalities were defined as bilateral NPi < 3. Diff Size and Diff NPi were considered unilateral pupil abnormalities since one abnormal pupil would be sufficient to make pupils asymmetric between eyes.

We also explored incidence, defined dichotomously as present or absent if a pupil abnormality ever occurred during the first 72 hours in a patient. Sensitivity analyses including only NPi < 3, Diff NPi > 0.7 or size > 1mm were also performed (**eMethods**).

### Analysis

Baseline cohort characteristics were summarized for the full cohort and stratified by TBI severity group with categorical variables reported as frequency (proportion) and continuous variables reported as median (25^th^ – 75^th^ percentile) as shown in Table 1. When reporting pupil abnormality frequencies, we only included frequencies for patients with at least one pupil abnormality (**Table 2**). To compare the incidence and frequency of pupil abnormalities between different TBI severity groups, we used the chi-square test and Fisher’s exact test for categorical variables. For continuous variables, we employed ANOVA and the Kruskal-Wallis test (**eMethods**).

**Table 1.**
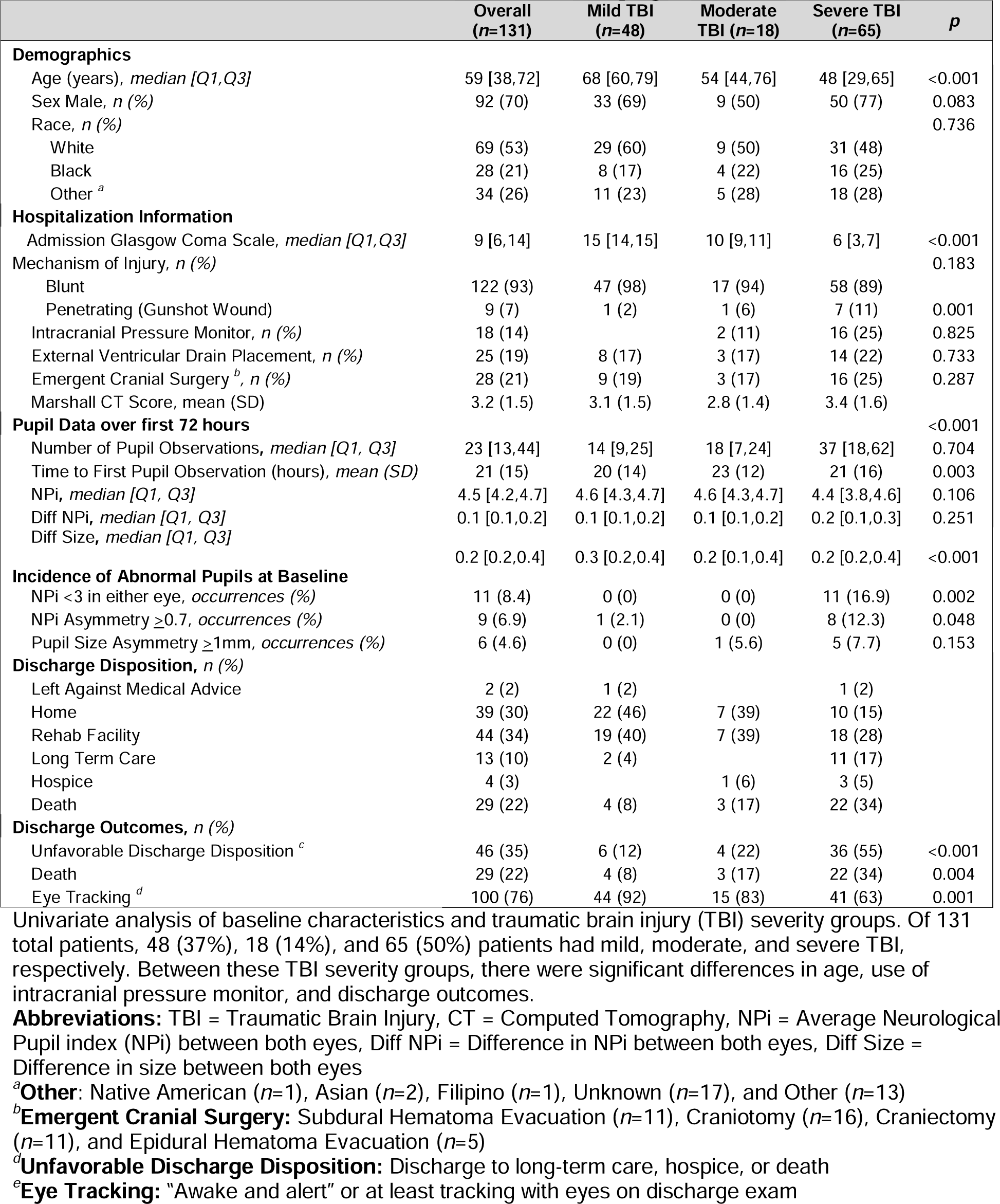
Baseline Characteristics of Traumatic Brain Injury Patients (*n* = 131)

**Table 2.** Incidence and Frequency of Pupil Abnormalities over first 72 hours in Traumatic Brain Injury Patients (*n* = 131)

| | Overall<br>($n=131$) | Mild TBI<br>($n=48$) | Moderate<br>TBI ($n=18$) | Severe<br>TBI ($n=65$) | $p$ |
| --- | --- | --- | --- | --- | --- |
| **Incidence, $n$ (%)** |  |  |  |  |  |
| Pupil Abnormalities <sup>a</sup> | 98 (75) | 30 (62) | 11 (61) | 57 (88) | 0.002 |
| Unilateral Pupil Abnormalities <sup>b</sup> | 95 (73) | 29 (60) | 11 (61) | 55 (85) | 0.008 |
| Bilateral Pupil Abnormalities <sup>c</sup> | 24 (18) | 3 (6) | 1 (6) | 20 (31) | 0.001 |
| **Frequency, median [Q1, Q3] <sup>d</sup>** |  |  |  |  |  |
| Pupil Abnormalities <sup>a</sup> | 18 [10,40] | 17 [6,24] | 30 [12,43] | 20 [12,55] | <0.001 |
| Unilateral Pupil Abnormalities <sup>b</sup> | 17 [8,29] | 17 [7,25] | 30 [11,43] | 16 [10,32] | 0.028 |
| Bilateral Pupil Abnormalities <sup>c</sup> | 15 [7, 92] | 9 [7,13] | 4 [4,4] | 15 [7,100] | N/A <sup>e</sup> |

Univariate analysis of incidence and frequency characteristics over first 72 hours and traumatic brain injury (TBI) severity groups. Incidence and frequency of all pupil abnormalities differed significantly among TBI severity groups. Frequency is reported as percentage and includes only patients with nonzero frequencies.
**Abbreviations:** TBI = Traumatic Brain Injury
<sup>a</sup>**Pupil Abnormalities:** Neurological Pupil index (NPi) < 3, NPi difference between both eyes (Diff NPi) $\geq 0.7$ , or Size difference between both eyes (Diff Size) $\geq 1$ mm
<sup>b</sup>**Unilateral Pupil Abnormalities:** Unilateral NPi < 3, Diff NPi $\geq 0.7$ , or Diff Size $\geq 1$ mm
<sup>c</sup>**Bilateral Pupil Abnormalities:** Bilateral NPi < 3
<sup>d</sup>**Frequency:** frequency was calculated only for patients with nonzero frequencies
<sup>e</sup>**N/A:** unable to perform statistical test for $p$ value due to low sample size

To test our primary hypothesis, we performed multivariable logistic regression of pupil abnormality frequency and unfavorable discharge disposition after adjusting for hypothesized confounders including age, race, mechanism of injury, TBI severity, and Marshall CT score.^15,16^ We performed similar analyses using secondary exposures and outcomes and performed subgroup analysis of patients with severe TBI. Given that the data was not normally distributed, reporting between the median or mean was chosen based on sample size and distribution.

To determine the prognostic value of pupil abnormality frequency, we modeled unfavorable discharge disposition and death at discharge in moderate and severe TBI using the core IMPACT TBI model and IMPACT plus pupil abnormality frequency over the first 72 hours. Features of the base IMPACT model included age, the motor component of GCS on admission, and pupil reactivity on admission. Pupil reactivity was defined subjectively and dichotomously as either bilateral or unilateral pupil reactivity, with bilateral pupil reactivity as the reference. We compared model goodness-of-fit using the likelihood ratio test and evaluated model performance using receiver operating characteristic (ROC) curve (**eMethods**).^16^

Significance threshold of α=0.05 was used our primary hypothesis. All other tests were hypothesis generating only. Preprocessing and analysis of data were done using code publicly available at https://github.com/fordivyav/TBI_Pupillometry (**eMethods**). The study was approved by the Boston Medical Center Institutional Review Board. Informed consent was waived due to the anonymous nature of the study. Anonymized data for this study are available from the senior author (C.J.O.) upon reasonable request.

## Results

### Cohort Characteristics

Our final cohort consisted of 131 patients with 3877 total pupil measurements. Thirty percent of the cohort was female. Median age was 59 years [38, 72] (25^th^, 75^th^ percentile). Thirty-seven percent (n=48) of patients had mild TBI, 14% (n=18) had moderate TBI, and 50% (n=65) had severe TBI. Seven percent (n=9) of the cohort had penetrating injuries. Median Marshall CT score was 2 [2, 5]. Unfavorable discharge disposition occurred in 35% (n=46) of patients, with 22% (n=29) of patients deceased upon discharge, with 4.6% of the total cohort who died prior to 72 hours (**Table 1**). Patients underwent a median of 23 [13, 44] pupil measurements. Median time to first pupil measurement was 3 hours [2, 6]. Median NPi over 72 hours was 4.4 [4.1, 4.6]. Seventy-five percent (n=98) of TBI patients experienced at least one pupil abnormality. Among those patients with pupil abnormality, median frequency was 18% [10, 40]. Of patients with at least one abnormal pupil measurement, 97% (n=95) of patients experienced unilateral pupil abnormalities, and 24% (n=24) experienced bilateral pupil abnormalities. Eighteen percent (n=18) had occurences of both unilateral only and bilateral pupil abnormalities.

Median frequency of unilateral and bilateral pupil abnormalities was 17% [8-29], and 15% [7-92%] respectively (**Table 2**, **Figure 1, eTable 1**). Patients with unfavorable discharge disposition had increased median frequency of pupil abnormalities compared to patients with favorable discharge disposition (26 [8, 70] vs. 8% [0, 20]) (**eFigure 2**).

**Figure 2:**
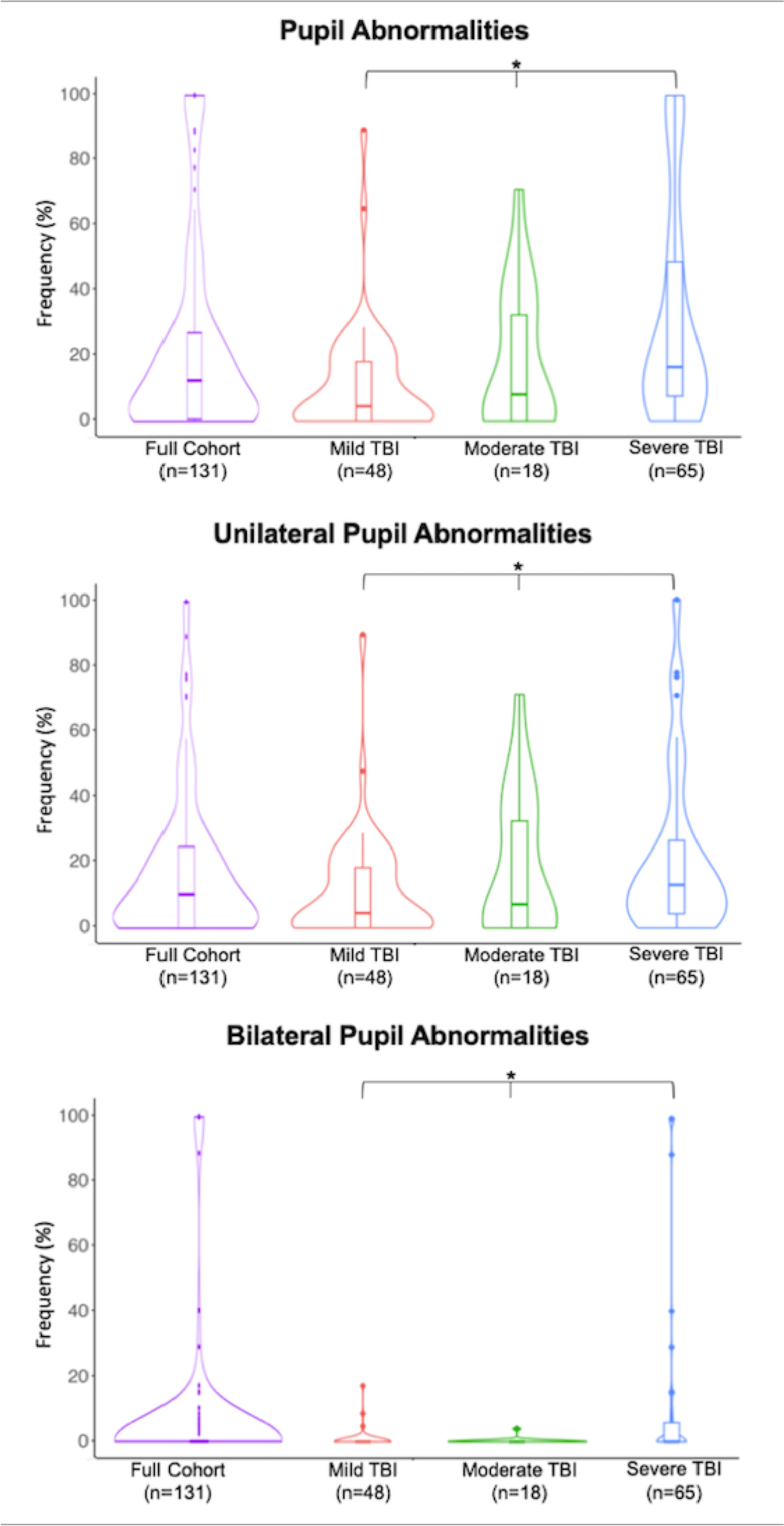
Violin Plots of Pupil Abnormality Frequency by TBI Severity. Pupil abnormalities including unilateral and bilateral pupil abnormalities differed significantly across all TBI severity groups. Frequencies were reported as percentages and only include patients with nonzero frequencies. Pupil Abnormalities and unilateral pupil abnormalities occurred in all TBI severity groups, with increased median frequency associated with increased TBI severity. In contrast, bilateral pupil abnormalities were rare in the full cohort with most bilateral Pupil Abnormalities occurring in severe TBI. **Abbreviations:** % = Percent, TBI = Traumatic Brain Injury, * = statistically significant difference across TBI severity groups (*p* < 0.05)

### Pupil Abnormalities across TBI Severity Groups

There were significant differences observed in age, intracranial pressure monitor use, and discharge outcomes among TBI severity groups (**Table 1**). Patients with severe TBI were more likely to experience at least one pupil abnormality (88% severe vs. 62% mild vs. 61% moderate) (*p*=0.002) (**eFigure 4**). Bilateral pupil abnormality incidence was much higher in severe TBI (31%) compared to mild and moderate TBI (6% each) (*p*=0.001) (**Figures 1-2**).

In patients who experienced pupil abnormalities (n=98), the frequency differed across TBI severity groups as follows: 20% [12, 55] in severe, 30% [12, 43] in moderate, and 18% [10, 40] in mild (*p*<0.001) (**Table 2, eFigures 3-4**).

**Figure 3:**
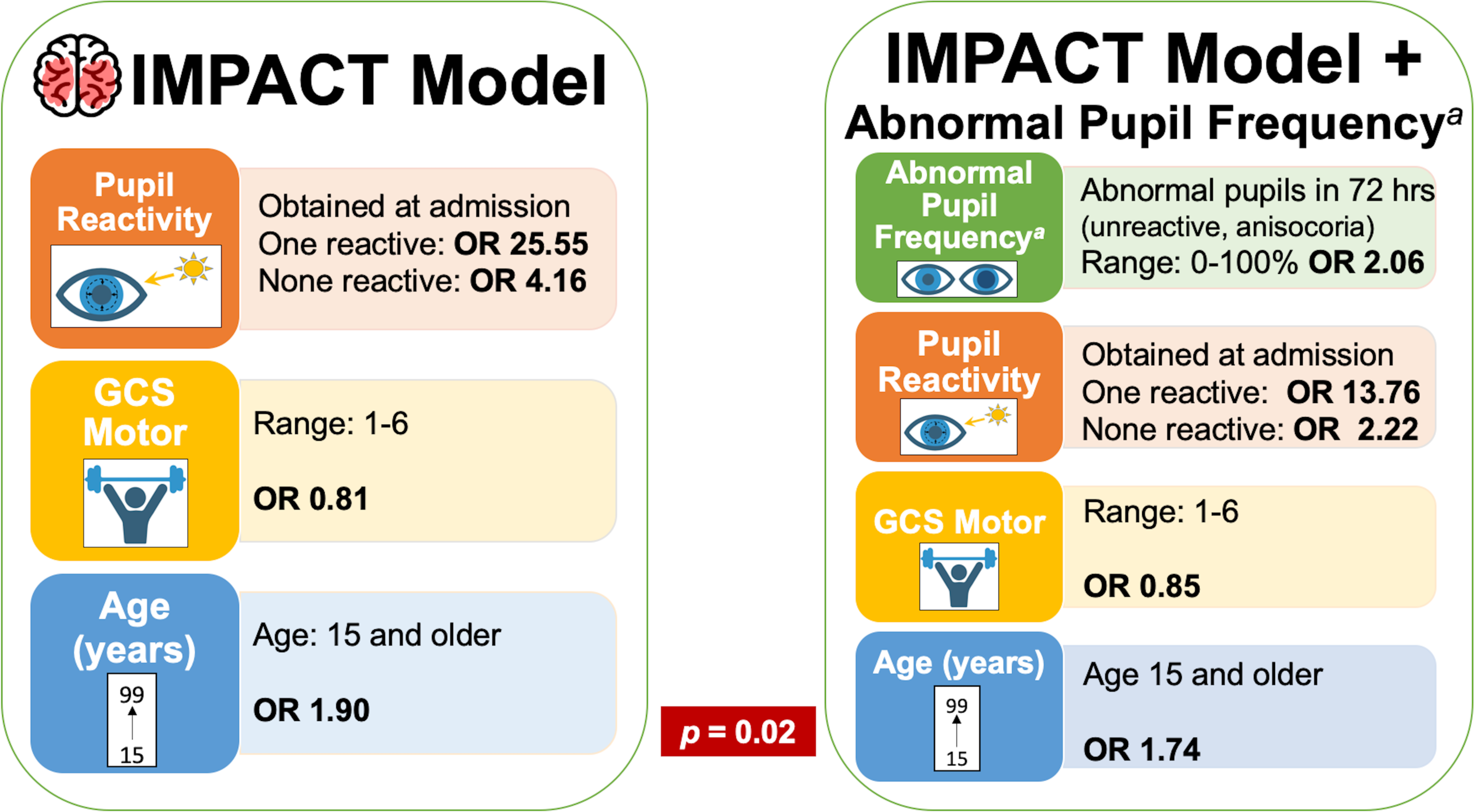
Schematic for IMPACT Model with and without Abnormal Pupil Frequency. Created with BioRender.com. The IMPACT model consisted of pupil reactivity on admission (both, one, or neither reactive), Glasgow Coma Scale motor component on admission, and age trained on our study cohort. The IMPACT model with pupil abnormality frequency over the first 72 hours of admission had improved goodness-of-fit (*p* = 0.02) compared to the IMPACT model alone. **Abbreviations:** IMPACT Model = IMPACT TBI Model, TBI = Traumatic Brain Injury, OR = Odds Ratio, GCS = Glasgow Coma Scale, Abnormal Pupil Frequency = Pupil Abnormality Frequency, Hrs = hours *^a^* **Abnormal Pupil Frequency**: Pupil abnormality frequency over the first 72 hours of admission (Neurological Pupil index [NPi] < 3, NPi difference between both eyes ≥ 0.7, or Size difference between both eyes > 1mm)

To gain insight into the relation between pupil abnormalities in patients without severe TBI, we describe the hospital courses for mild and moderate TBI patients with NPi < 3 (**eTable 2**).

### Pupil Abnormality Frequency and Discharge Dispostion

In our adjusted multivariable model, we found that a 1% increase in pupil abnormality frequency was associated with unfavorable discharge disposition (OR: 1.03; 95% Cl, 1.01-1.05). We observed that the OR increased in our stratified analysis of bilateral pupil abnormality frequency (OR: 1.22; 95% Cl, 1.07-1.45), but not in unilateral pupil abnormality frequency (OR: 1.02; 95% Cl, 1.00-1.04) (**Table 3**). Both bilateral and unilateral pupil abnormality frequency were associated with mortality (**Table 3**). Comprehensive results including subanalysis of our severe TBI subgroup and single pupil abnormality features are included in eTables 3-6.

**Table 3.** Multivariable Models of Pupil Abnormalities Frequency on Discharge Outcomes (*n* = 131)

| Frequency <sup>a</sup> | Unfavorable Discharge Disposition |  |  | Death at Discharge |  |  |
| --- | --- | --- | --- | --- | --- | --- |
|  | OR | 95% CI | p | OR | 95% CI | p |
| Pupil Abnormalities <sup>b</sup> | 1.03 | 1.01-1.05 | 0.002 | 1.05 | 1.03-1.07 | <0.001 |
| Unilateral Pupil Abnormalities <sup>c</sup> | 1.02 | 1-1.04 | 0.132 | 1.02 | 1-1.05 | 0.047 |
| Bilateral Pupil Abnormalities <sup>d</sup> | 1.22 | 1.07-1.45 | 0.008 | 1.23 | 1.08-1.45 | 0.005 |

Multivariable logistic regression models were adjusted for age, race, severity group of traumatic brain injury, Marshall computed tomography score, and penetrating vs. blunt trauma. Frequency was reported as percentage. Frequency of pupil abnormalities and bilateral pupil abnormalities were associated with unfavorable discharge disposition and death at discharge. Unilateral pupil abnormalities were associated with death.
**Abbreviations:** OR = Odds Ratio, CI = Confidence Interval, n = number of patients in full cohort with specified pupil abnormality, M = number of pupil measurements with specified pupil abnormality
<sup>a</sup>**Frequency:** calculated for every 1% increase in frequency
<sup>b</sup>**Pupil Abnormalities:** Neurological Pupil index (NPi) < 3, NPi difference between both eyes (Diff NPi) ≥ 0.7, or Size difference between both eyes (Diff Size) ≥ 1mm
<sup>c</sup>**Unilateral Pupil Abnormalities:** Unilateral NPi < 3, Diff NPi ≥ 0.7, or Diff Size ≥ 1mm
<sup>d</sup>**Bilateral Pupil Abnormalities:** Bilateral NPi < 3

### IMPACT Score

The likelihood ratio test revealed that the IMPACT model plus pupil abnormality frequency had an improved goodness-of-fit compared to the core IMPACT model alone when predicting unfavorable discharge disposition (*X*^2^=5.67, *p*=0.02) (**Figure 3, eTable 7**). The IMPACT model for death at discharge also showed improved goodness-of-fit with the addition of pupil abnormality frequency (*p*<0.01) (**eTable 7**). Absent bilateral pupil reactivity at baseline (/3=13.6) and pupil abnormality frequency (/3=2.06 per 1% increase) were the most important coefficients for predicting unfavorable discharge disposition and death at discharge (**eTable 7, eFigure 5**). While the ROC curve showed improved performance for the IMPACT model with pupil abnormality frequency (Area under Curve [AUC] 0.78) compared to the IMPACT model alone (AUC 0.75), the differences in AUC were not statistically significant for our primary outcome (*p*=0.2) (**eFigure 6**). Similar results were seen when predicting death at discharge using the IMPACT model with (AUC 0.89) and without pupil abnormality frequency (AUC 0.81) (*p*=0.056) (**eFigure 6**).

## Discussion

In this study, we found that increased frequency of abnormal pupil measurements within the first 72 hours of admission is associated with unfavorable discharge disposition and improves the IMPACT TBI model, which uses subjective pupil reactivity assessment at baseline. These findings are significant because longitudinal pupillometric information is not standardly used for prognostication or treatment decision-making but has the potential to improve care.

Our study reveals that abnormal pupil measurements over the first 72 hours are common among TBI patients admitted to the ICU (75%), especially in severe TBI (88%). While patients with severe TBI are more likely to experience at least one pupil abnormality, they remain common among moderate and mild TBI patients as well. Our observation that at least one NPi < 3 occurred in 42% of patients is consistent with results from a recent prospective study investigating the association of abnormal pupil reactivity in patients with heterogenous acute brain injury (40%).^18^ The difference in prevalence of unilateral (73%) and bilateral (18%) pupil abnormalities in patients who had at least one abnormal pupil measurement is most likely due to the more inclusive definition we used for unilateral pupil abnormalities including size difference and NPi difference.

In patients who experienced pupil abnormalities, the median frequency over the first 72 hours was higher in moderate and severe TBI patients compared to those with mild TBI. While moderate TBI patients had a non-statistically significant higher frequency of pupil abnormalities than severe TBI patients, we posit this may be because they experienced neurologic worsening events within the first 72 hours of admission (**eTable 2**), evidenced by the significant association between increased pupil abnormality frequency and discharge disposition.

Our review of mild and moderate TBI patients with NPi < 3 revealed that patients with mild TBI and pupil abnormalities were frequently complicated by intraparenchymal or subdural hemorrhages, some of which underwent surgical treatment (**eResults**). We infer that these cases support the hypothesis that pupil abnormality incidence and frequency signify increased severity that may not be reflected on admission. This finding is likely reflective that patients with milder TBI phenotypes at admission can deteriorate during the hospital course. Following quantitative pupillometry in moderate or even mild TBI patients may be important to identify neurologic worsening, which can occur in 35% of TBI patients within the first 24 hours of admission.^19^

Our findings regarding the association of pupil abnormality frequency and outcome are consistent with prior studies examining quantitative poor pupil reactivity over admission in heterogeneous causes of acute brain injury or in small case series of TBI^.16,20,21^ Our observation that the relation between pupil abnormality frequency and outcome remains significant for bilateral but not for unilateral pupil abnormalities may be due to the fact that the bilateral abnormalities can signify irreversible intrinsic damage to pathways for recovery in the midbrain and brainstem, whereas unilateral abnormalities may be more context dependent and reversible.

Our study demonstrates that the inclusion of pupil abnormality frequency over the first 72 hours of admission improves goodness-of-fit for current prognostic models. While the ROC curves for the IMPACT TBI models improved with the addition of pupil abnormality frequency for both outcomes, results were not statistically significant, most likely due to the limited number of moderate and severe TBI patients.

There are several study limitations. As a retrospective, observational study, we cannot elucidate any cause-effect relationship or exclude all potential confounding factors, but this serves as a preliminary analysis into the role of pupil abnormalities in TBI prognostication. We did not control for pain, ambient light, or cognitive load, which have been shown to affect pupil reactivity.^22-24^ While our study was single-center with a relatively small sample size, a post hoc analysis revealed we had a power of 98% with a sample size of 131 when detecting the observed difference in abnormal pupil frequency between patients with favorable and unfavorable outcomes. Additional tests for other exposures were not powered sufficiently and thus are hypothesis generating only. We had varying number of measurements per patient, which can lead to bias. Moreover, we acknowledge that early mortality or withdrawal of life-sustaining therapy prior to 72 hours may have confounded results, though this was a minority of our patients (4.6%).

While we conducted multiple tests of association, we selected only one primary hypothesis to minimize this risk of false-positive associations. We acknowledge that the IMPACT model was designed to detect unfavorable outcome and six-month mortality, and that we did not have access to long-term follow up to compare outcomes as originally designed.

Despite these limitations, our study had several strengths. We had access to over 3,000 unique pupil measurements obtained from a heterogeneous and diverse patient cohort. We performed subgroup analyses and explored clinically meaningful outcomes to better understand pupil abnormalities in TBI.

## Conclusions

Pupil abnormalities are common in TBI patients over the first 72 hours of injury, especially in those with severe TBI. Increased pupil abnormality frequency may be associated with higher risk of unfavorable discharge disposition, and pupil abnormality inclusion improves the IMPACT TBI model for prognostication. Future studies should be conducted to further evaluate and externally validate whether the inclusion of pupil abnormality frequency in current models improves prognostication and treatment decisions.

## Author contributions

Dr. Ong and Dr. Veerapeneni designed the study. So Yeon Kim was involved in data acquisition. Dr. Veerapeneni was involved in data analsyis and interpretation. Dr. Veerapeneni, Naveen Arunachalam Sakthiyendran, Dr. Mohammed, Dr. Dupuis, and Dr. Ong were involved in drafting the original manuscript. Dr. Veerapeneni, Naveen Arunachalam Sakthiyendran, Dr. Daneshmand, Dr. Sheth, Dr. Abdalkader, Dr. Greer, Dr. Mohammed, Dr. Gilmore, Dr. Dupuis, and Dr. Ong were involved in critical revisions. Dr. Daneshmand and Dr. Abdlakdaer were involved in validation. Dr. Veerapaneni and Chuong Nguyen helped with data analysis software. So Yeon Kim helped with project administrative tasks and resources. Dr. Ong helped supervise and aquire funding for the project.

## Data Availability

All data produced in the present study are available upon reasonable request to the authors

## Acknowledgements

We would like to thank the neurocritical care nurses at Boston Medical Center for their valuable contribution of pupillometric data to this study. Additionally, we thank Linda Rosen from the Boston University Clinical Data Warehouse for helping to procure data from electronic health records.

## Statement of Human Rights

This study is adherent to ethical guidelines and approved by the Boston Medical Center Institutional Review Board. For this type of study, formal consent is not required.

## Supplementary Appendix for

### eMethods: Data Collection and Preprocessing of Pupillometry Data

#### Data Collection

All extracted demographic and imaging data were maintained in REDCap (Research Electronic Data Capture)^1^, a secure, web-based software platform designed to support data capture for research studies. Race, admission Glasgow Coma Scale, mechanism of trauma (blunt vs. penetrating), emergent cranial surgery, use of intracranial pressure monitor, external ventricular drain placement, Marshall computed tomography (CT) score, presence of exclusion criteria, death at discharge, and eye tracking on discharge were obtained through chart review. Details for criteria used for classification are below:

| Variable | Criteria |
| --- | --- |
| Race (white vs. black vs. other) | Reference race as White.<br>Patients were considered “other” if American Indian, Asian, Filipino race, or unknown. |
| Admission Glasgow Coma Scale | Per chart review, if patient had a Glasgow Coma Scale (GCS) that changed within the first 6 hours of admission, the GCS and severity of traumatic brain injury based on the GCS was adjusted to the more recent value.<br>Mild TBI = GCS 13-15<br>Moderate TBI = GCS 9-12<br>Severe TBI = GCS 3-8 |
| Mechanism of Trauma (blunt vs. penetrating) | Patients were classified into penetrating vs. blunt mechanism of trauma based on if they had a gunshot wound (“penetrating”) vs. other cause of trauma (“blunt”). |
| Emergent Cranial Surgery | Patient having a craniectomy, craniotomy, or evacuation of a hematoma within the first 72 hours. |
| Marshall Computed Tomography (CT) Score | Diffuse injury V was classified first if a patient had decompressive surgery for traumatic brain injury within the first 72 hours of admission.<br>Otherwise, Marshall CT score was calculated by direct evaluation of the most severe CT brain imaging taken within the first 72 hours and associated imaging radiology report. |
| Discharge Disposition | Patient discharge disposition was classified as “Rehab Facility” for both “acute rehab facility” and “skilled nursing facility”. |

To determine reliability of the Marshall CT scores arbitrated by D.V., we randomly selected 10% of patients and gave their corresponding brain images to a second trained researcher (A.D.) for re-evaluation of the Marshall score. Inter-rater reliability was computed by comparing CT scores from D.V. and A.D. and found to be 70%. Discrepant features were reviewed by D.V. and A.D. to establish consensus.

### eMethods: Exposures

We characterized pupil abnormality as the following for further exploratory analysis:

#### Isolated Features of Pupil Abnormalities

1. NPi < 3
2. Diff NPi > 0.7
3. Diff Size ≥ 1mm

All pupil abnormalities are inclusive. In addition to frequency, incidence, and continuous variables of NPi, Diff NPi, and Diff Size were also examined as part of exploratory analysis.

### eMethods: Outcomes

Our exploratory outcome was eye tracking on discharge. This was defined as “awake or alert” or able to track objects on discharge exam. We chose this as an exploratory outcome because conscientiousness is an important clinical factor to patients and their families, especially relating to goals of care discussions.

### eMethods: IMPACT Models

To determine whether pupillometry over 72 hours improves current prognostic TBI models, we used the three parameters from the core IMPACT model: age, motor component of admission Glasgow Coma Scale, and pupil reactivity on admission (none, unilateral, or bilateral reactivity) to predict both unfavorable discharge disposition and death at discharge. All 3 input factors were obtained through chart review. All input factors had single values for each patient. Pupil reactivity was at baseline. Input parameters were then normalized including pupil abnormality frequency. Pupil abnormality frequency was a single measurement for each patient.

The IMPACT model was created by training a logistic regression model to predict unfavorable discharge disposition on our whole cohort using the normalized input parameters. Then the combined IMPACT and pupil frequency model was created using the same method except with the addition of the normalized pupil abnormality frequency input parameter. We used the likelihood ratio test to evaluate goodness-of-fit between the two models for both outcomes (unfavorable discharge disposition and death at discharge). In addition, we also created a receiver operating characteristic (ROC) curve to compare the IMPACT model with and without abnormal pupil frequency for both outcomes.

Our R pseudocode for the likelihood ratio test was the following:

- IMPACT model = glm(Outcome ∼ Age + Motor Component of Glasgow Coma Scale + Pupil Reactivity on admission, data = study cohort)
- IMPACT + Abnormal Pupil Frequency model = glm(Outcome ∼ Age + Motor Component of Glasgow Coma Scale + Pupil Reactivity on admission + Pupil Abnormalities Frequency over 72 hours, data = study cohort).
- lrtest(IMPACT model, IMPACT + Abnormal Pupil Frequency model)

### eMethods: Preprocessing of Data and Statistical Analyses

#### Computer Programming Languages

Preprocessing of data and statistical analyses was done using Python Version 3.9.13 (Van Rossum, G., & Drake, F. L. (2009). *Python 3 Reference Manual*. Scotts Valley, CA: CreateSpace) and R Version 4.2.0 (R Core Team (2023). R: A language and environment for statistical computing. R Foundation for Statistical Computing, Vienna, Austria).

#### Preprocessing of Pupillometry Data

Pupillometry data was preprocessed using both Python Version 3.9.13 (Van Rossum, G., & Drake, F. L. (2009). *Python 3 Reference Manual*. Scotts Valley, CA: CreateSpace) and R Version 4.2.0 (R Core Team (2023). R: A language and environment for statistical computing. R Foundation for Statistical Computing, Vienna, Austria) and is publicly available at https://github.com/fordivyav/TBI_Pupillometry. Pupil measurements were preprocessed as follows:

For all pupil observations during a hospital admission, we calculated the minimum NPI, NPI difference, and size difference between both the left and right eyes for the patient. We defined the “start of hospitalization” for each patient as either the admission time (if precedes the time of first pupil measurement) or else the presentation time. We then filtered for pupil data up to 72 hours after the “start of hospitalization.” Patients with less than three pupil observations within the first 72 hours were excluded from our cohort.

To create the final dataset, we took the 72-hour dataset for all patients and computed the median NPi, minimum NPi, NPi difference, and size difference for each patient. We computed pupil abnormality frequencies by calculated the number of pupil measurements in which the pupil abnormality was present divided by the total number of pupil measurements in the 72-hour dataset for the patient. Incidence for each pupil abnormality was then computed at the patient level. Thus, incidence and frequency of pupil abnormality was consolidated as a single value per patient using data from pupil measurements up to 72 hours after admission.

We used the python package “tableone” ^2^ to perform the Chi square or ANOVA test on demographic information, clinical data, and pupil data across the TBI severity groups. For categorical variables in which the expected value for any cell was less than five for the Chi square test, we used the Fisher’s exact test instead. Similarly, for frequency of pupil abnormalities we used the Kruskal-Wallis test to account for the zero-inflated distributions. Because frequency had a zero-inflated distribution, we also reported frequency (%) as median [Q1, Q3] of only non-zero frequency values.

For post-hoc power analyses, we performed the Wilcoxon-Mann-Whitney test in R for both primary and secondary exposures and our primary outcome.

### eResults: Descriptive Analysis of Hospital Course

We conducted descriptive analysis of hospitalization course for patients with mild or moderate TBI who have NPi < 3. Of eleven mild TBI patients with NPi < 3, the only patient who died had an intraparenchymal hemorrhage with bilateral NPi < 3, diff NPi > 0.7, and diff Size ≥ 1. Of mild TBI patients who had bilateral NPi < 3, one had intraparenchymal hemorrhage, one had subdural hematoma vs. epidural hematoma, and one had subdural hematoma, intraparenchymal hemorrhage, and frontotemporal hematoma, of which one expired, one underwent a craniotomy, and the other improved and was discharged home after 4 days, respectively. Only one patient with a subdural hematoma, intraparenchymal hemorrhage, and subarachnoid hemorrhage after a motor vehicle accident who underwent emergent craniotomy was discharged to long term care. Out of the three moderate TBI patients with NPi < 3, none had unfavorable discharge disposition. Only one had bilateral NPi < 3 due to a gunshot wound, underwent craniectemy, and was discharged to rehab after 71 days. The rest were discharged to rehab.

**eTable 1:** Incidence and Frequency of Pupil Abnormalities over first 72 hours in Traumatic Brain Injury Patients.

|  | Overall<br>(n=131) | Mild TBI<br>(n=48) | Moderate<br>TBI (n=18) | Severe<br>TBI (n=65) | *p* |
| --- | --- | --- | --- | --- | --- |
| **Incidence among whole cohort, *n* (%)** |  |  |  |  |  |
| Pupil Abnormalities <sup>a</sup> | 98 (75) | 30 (62) | 11 (61) | 57 (88) | 0.002 |
| NPi < 3 | 55 (42) | 12 (25) | 3 (17) | 40 (62) | <0.001 |
| Diff NPi ≥ 0.7 | 72 (55) | 14 (29) | 9 (50) | 49 (75) | <0.001 |
| Diff Size ≥ 1 | 74 (56) | 24 (50) | 8 (44) | 42 (65) | 0.163 |
| Unilateral Pupil Abnormalities <sup>b</sup> | 95 (73) | 29 (60) | 11 (61) | 55 (85) | 0.008 |
| Bilateral Pupil Abnormalities <sup>c</sup> | 24 (18) | 3 (6) | 1 (6) | 20 (31) | 0.001 |
| **Frequency among whole cohort, median [Q1,Q3]** |  |  |  |  |  |
| Pupil Abnormalities <sup>a</sup> (m = 30) | 18 [10,40] | 17 [6,24] | 30 [12,43] | 20 [12,55] | <0.001 |
| NPi < 3 (m = 38) | 14 [5,40] | 8 [4,17] | 8 [7,12] | 15 [6,60] | <0.001 |
| Diff NPi ≥ 0.7 (m = 38) | 11 [5,22] | 17 [12,24] | 12 [4,13] | 9 [5,22] | <0.001 |
| Diff Size ≥ 1 (m = 30) | 11 [6,23] | 9 [5,17] | 26 [16,34] | 12 [6,25] | 0.168 |
| Unilateral Pupil Abnormalities <sup>b</sup> (m = 30) | 17 [8,29] | 17 [7,25] | 30 [11,43] | 16 [10,32] | 0.028 |
| Bilateral Pupil Abnormalities <sup>c</sup> (m = 36) | 15 [7, 92] | 9 [7,13] | 4 [4,4] | 15 [7,100] | N/A<sup>d</sup> |

Univariate analysis of incidence and frequency characteristics over 72 hours and TBI severity groups. Both incidence and frequency of primary exposures (any and unilateral pupil abnormalities) differ significantly across the TBI severity groups. *P* values were calculated using analysis of variance tests.
**Abbreviations:** TBI = Traumatic Brain Injury, NPi = Average Neurological Pupil index (NPi) between both eyes, Diff NPi = Difference in NPi between both eyes, Diff Size = Difference in size between both eyes, m = median number of pupil measurements per patient for patients with nonzero frequency
<sup>a</sup>**Pupil Abnormalities:** NPi < 3, Diff NPi ≥ 0.7, or Diff Size ≥ 1mm
<sup>b</sup>**Unilateral Pupil Abnormalities:** Unilateral NPi < 3, Diff NPi ≥ 0.7, or Diff Size ≥ 1mm
<sup>c</sup>**Bilateral Pupil Abnormalities:** Bilateral NPi < 3

**eTable 2:** Hospital Course for Mild and Moderate Traumatic Brain Injury Patients with NPi < 3.

| Mild Traumatic Brain Injury |  |  |  |  |  |  |
| --- | --- | --- | --- | --- | --- | --- |
|  | Pupil Abnormalities | GCS | Length of Stay (days) | Imaging Findings | Outcome | Summary of Hospital Course |
| **Patient 1** | Bilateral NPi < 3<br>Diff NPi ≥ 0.7<br>Diff Size ≥ 1 | 14 | 4 | 9mm Intraparenchymal Hemorrhage | Deceased | Elderly male with stage 4 bladder cancer, dementia, chronic obstructive pulmonary disease, and deep vein thrombosis on anticoagulation presented with fall. Found to have intraparenchymal hemorrhage. Made comfort measures only day 2 and died day 4. |
| **Patient 2** | Unilateral NPi < 3<br>Diff NPi ≥ 0.7 | 14 | 6 | Acute on Chronic Right Subdural Hematoma, Subarachnoid Hemorrhage | Home | Middle-aged male with polysubstance use disorder and schizoaffective disorder with multiple falls presented as a code stroke with possible assault and subdural hematoma. Improved and discharged home day 6. |
| **Patient 3** | Bilateral NPi < 3<br>Diff NPi ≥ 0.7<br>Diff Size ≥ 1 | 13 | 4 | Right Subdural Hematoma, Intraparenchymal Hemorrhage x 3, Frontotemporal Hematoma | Home | Elderly male with dementia, stroke, and seizure disorder presents after fall with confusion. Found to have subdural hematoma, intraparenchymal hemorrhage and frontotemporal hematoma. Improved and discharged home day 4. |
| **Patient 4** | Unilateral NPi < 3<br>Diff NPi ≥ 0.7 | 15 | 7 | 16mm Right Subdural Hematoma with 6mm Midline Shift | Rehab Facility | Elderly female with breast cancer and many falls presents with fall. Found to have subdural hematoma. Treated with hyperosmolar therapy and discharged to rehab day 7. |
| **Patient 5** | Bilateral NPi < 3 | 13 | 4 | Acute on Chronic Subdural Hematoma vs. Epidural Hematoma | Home | Middle-aged male with alcohol use disorder presented intoxicated after rolling on broken dishes on floor. Found to have frontoparietal hematoma. Underwent craniotomy day 1. Discharged home day 4. |
| **Patient 6** | Unilateral NPi < 3<br>Diff NPi ≥ 0.7<br>Diff Size ≥ 1 | 14 | 4 | Acute on Chronic Bilateral Subdural Hematoma | Skilled Nursing Facility | Elderly male with dementia, cirrhosis, chronic kidney disease, diabetes, atrial fibrillation, and anemia presented with fall. Found to have subdural hematoma. Underwent bilateral middle meningeal embolization and discharged to skilled nursing facility day 4. |
| **Patient 7** | Unilateral NPi < 3<br>Diff NPi ≥ 0.7<br>Diff Size ≥ 1 | 14 | 13 | Acute 1cm Right Subdural Hematoma with 7mm Midline Shift, Possible Right Transverse and Sigmoid Sinus Thrombus | Long Term Care | Male in his 60s with HIV, hypertension, and hepatitis presented with 2-3 days of left sided weakness, slurred speech, and lethargy 2 weeks after admission for fall with TBI. Found to have subdural hematoma and underwent craniotomy day 1. Course complicated by concern for seizure with new subarachnoid hemorrhage and possible sinus thrombosis. Discharged to long term care day 13. |
| **Patient 8** | Unilateral NPi < 3<br>Diff NPi ≥ 0.7<br>Diff Size ≥ 1 | 14 | 5 | Right Corona Radiata and Left Subarachnoid Hemorrhage | Home | Middle-aged male with hypertension, polysubstance use disorder, hepatitis C, and TBI presents after motor vehicle accident while intoxicated. Found to have subarachnoid hemorrhage with repeat head CT showing increase but later stabilized. Patient advised to go to acute rehab, but preferred home. Discharged home day 5. |
| **Patient 9** | Unilateral NPi < 3<br>Diff Size ≥ 1 | 15 | 20 | 4.5mm Subdural Hematoma and Right Frontal Lobe Periventricular Hypodensity | Skilled Nursing Facility | Elderly male with congestive heart failure, hypertension, diabetes, and multiple strokes presents with left upper extremity numbness after falling downstairs. Found to have subdural hematoma requiring craniotomy day 1. Course complicated by seizure, pneumonia, and pleural effusion. Discharged to SNF day 20. |
| **Patient 10** | Unilateral NPi < 3<br>Diff NPi ≥ 0.7<br>Diff Size ≥ 1 | 15 | 99 | Left Subdural Hematoma with 4mm Midline Shift, Intraparenchymal Hemorrhage of Right Posterior Temporal Lobe | Rehab Facility | Elderly male with history of hypertension, renal cell carcinoma, childhood motor vehicle collision requiring cranial plating, seizures, chronic lymphocytic leukemia, carotid stenosis on aspirin and Plavix presents after a fall with TBI. Found to have subdural hematoma and intraparenchymal hemorrhage requiring transfusion. Course complicated by cardiac arrest, complete heart block, acute hypoxemic respiratory failure, and subglottic stenosis. Multiple nodules noted concerning for metastatic cancer. Discharged to rehab day 99. |
| **Patient 11** | Unilateral NPi < 3<br>Diff NPi ≥ 0.7 | 15 | 10 | Subarachnoid Hemorrhage, 9mm Subdural Hematoma, Expanding Contusions | Skilled Nursing Facility | Male in his 60s with alcohol use disorder, stroke on anticoagulation, hypertension, and pancreatitis presents with fall. Found to have subarachnoid hemorrhage, subdural hematoma with expanding contusions, and carotid stenosis. Discharged to skilled nursing facility day 10. |
| **Moderate Traumatic Brain Injury** |  |  |  |  |  |  |
|  | **Pupil Abnormalities** | **GCS** | **Length of Stay (days)** | **Imaging Findings** | **Outcome** | **Summary of Hospital Course** |
| **Patient 1** | NPi < 3<br>Diff NPi ≥ 0.7 | 10 | 20 | 1.5cm Right Subdural Hematoma with 7mm Midline Shift | Skilled Nursing Facility | Elderly female with baseline dementia and osteopenia presented after fall with confusion. Found to have subdural hematoma requiring craniotomy day 1. Due to dementia, discharge to skilled nursing facility delayed to day 20. |
| **Patient 2** | Bilateral NPi < 3<br>Unilateral NPi < 3<br>Diff NPi ≥ 0.7<br>Diff Size ≥ 1 | 10 | 71 | Bullet Fragments in Right Parietal Region with Gas, Ballistic Wound with Multiple Bone Fragments, Small Right Subarachnoid Hemorrhage | Rehab Facility | Female in her 30s presented with a gunshot wound to right temple. Arrested in trauma bay and CT scanner. Day 2 underwent craniectomy and wound debridement. Course complicated by likely stress-induced cardiomyopathy, acute hypoxemic respiratory failure, and deep vein thrombosis. Discharged to rehab day 71. |
| **Patient 3** | Unilateral NPi < 3<br>Diff NPi ≥ 0.7<br>Diff Size ≥ 1 | 10 | 12 | Bilateral Frontal Hemorrhagic Contusions | Rehab Facility | Young male presented as an unrestrained vs. restrained driver in a head-on motor vehicle collision. Found to have bilateral frontal hemorrhagic contusions. Underwent multiple surgical repairs for trauma and discharged to rehab day 12. |

Eleven patients with mild TBI and 3 patients with moderate TBI had mean NPi across both eyes < 3. Out of patients with NPi < 3, two of the 11 mild patients had unfavorable discharge disposition, while none of the moderate TBI patients had an unfavorable discharge disposition.
**Abbreviations:** GCS = Glasgow coma scale on admission, TBI = Traumatic Brain Injury, NPi = Neurological Pupil index, Diff NPi = Difference in NPi between both eyes, Diff Size = Difference in size between both eyes

**eTable 3:** Subgroup Analysis of Patients with Severe Traumatic Brain Injury in first 72 hours.

|  | Primary Outcome |  |  | Secondary Outcome |  |  |  |  |  |
| --- | --- | --- | --- | --- | --- | --- | --- | --- | --- |
|  | Unfavorable Discharge Disposition |  |  | Death at Discharge |  |  | Eye Tracking |  |  |
| Continuous Variables | OR | 95%CI | p | OR | 95%CI | p | OR | 95%CI | p |
| NPi | 0.12 | 0.02-0.54 | 0.025 | 0.03 | 0-0.23 | 0.011 | 9.28 | 2.46-88.67 | 0.023 |
| Diff NPi | 2.64 | 0.67-18.36 | 0.234 | 2.65 | 0.7-14.79 | 0.203 | 0.42 | 0.07-1.58 | 0.258 |
| Diff Size | 1.5 | 0.3-9.73 | 0.637 | 6.73 | 1.02-71.26 | 0.076 | 0.25 | 0.03-1.56 | 0.174 |
| Incidence Variables |  |  |  |  |  |  |  |  |  |
| Pupil Abnormalities <sup>a</sup> | 1.41 | 0.26-8.86 | 0.693 | 4.64 | 0.42-136.48 | 0.273 | 0.13 | 0-1.51 | 0.158 |
| Unilateral Pupil Abnormalities <sup>b</sup> | 0.77 | 0.16-3.56 | 0.739 | 0.56 | 0.09-3.92 | 0.533 | 1.16 | 0.16-6.74 | 0.873 |
| Bilateral Pupil Abnormalities <sup>c</sup> | 8.11 | 1.85-48.98 | 0.01 | 72.72 | 7.83-2006.2 | 0.002 | 0.04 | 0-0.22 | 0.001 |
| Frequency Variables |  |  |  |  |  |  |  |  |  |
| Pupil Abnormalities <sup>a</sup> | 1.03 | 1.01-1.05 | 0.016 | 1.06 | 1.03-1.1 | 0.001 | 0.95 | 0.92-0.98 | 0.001 |
| Unilateral Pupil Abnormalities <sup>b</sup> | 1.01 | 0.99-1.04 | 0.371 | 1.02 | 0.99-1.04 | 0.185 | 0.98 | 0.96-1.01 | 0.189 |
| Bilateral Pupil Abnormalities <sup>c</sup> | 1.17 | 1.03-1.48 | 0.112 | 1.27 | 1.06-1.71 | 0.059 | 0.86 | 0.69-0.96 | 0.108 |

Multivariable logistic regression models were adjusted for age, race, Marshall computed tomography score, and penetrating vs. blunt trauma. In severe TBI, frequency of pupil abnormality was associated with unfavorable discharge disposition and death at discharge, while inversely associated with eye tracking.
**Abbreviations:** OR = Odds Ratio, CI = Confidence Interval, NPi = Average Neurological Pupil index (NPi) between both eyes, Diff NPi = Difference in NPi between both eyes, Diff Size = Difference in size between both eyes
<sup>a</sup>**Pupil Abnormalities:** NPi < 3, Diff NPi ≥ 0.7, or Diff Size ≥ 1mm
<sup>b</sup>**Unilateral Pupil Abnormalities:** Unilateral NPi < 3, Diff NPi ≥ 0.7, or Diff Size ≥ 1mm
<sup>c</sup>**Bilateral Pupil Abnormalities:** Bilateral NPi < 3

**eTable 4:** Multivariable Models of Median Continuous Pupil Values over first 72 hours and Discharge Outcomes.

|  | Primary Outcome |  |  | Secondary Outcome |  |  |  |  |  |
| --- | --- | --- | --- | --- | --- | --- | --- | --- | --- |
|  | Unfavorable Discharge Disposition |  |  | Death at Discharge |  |  | Eye Tracking |  |  |
| Continuous Variables | OR | 95%CI | p | OR | 95%CI | p | OR | 95%CI | p |
| NPi | 0.18 | 0.04-0.54 | 0.011 | 0.13 | 0.03-0.38 | 0.004 | 5.88 | 2.25-25.5 | 0.007 |
| Diff NPi | 2.26 | 0.67-13.16 | 0.277 | 2.23 | 0.76-9.73 | 0.204 | 0.45 | 0.10-1.35 | 0.221 |
| Diff Size | 2.60 | 0.6-14 | 0.231 | 13.14 | 2.35-102.0 | 0.007 | 0.12 | 0.02-0.63 | 0.02 |

Multivariable logistic regression models were adjusted for age, race, severity group of traumatic brain injury, Marshall computed tomography score, and penetrating vs. blunt trauma. NPi was associated with all outcomes. Diff Size was associated with death and inversely associated with eye tracking.
**Abbreviations:** OR = Odds Ratio, CI = Confidence Interval, NPi = Average Neurological Pupil index (NPi) between both eyes, Diff NPi = Difference in NPi between both eyes, Diff Size = Difference in size between both eyes

**eTable 5:** Multivariable Models of Pupil Abnormality Incidence within first 72 hours and Discharge Outcomes.

|  | Primary Outcome |  |  | Secondary Outcome |  |  |  |  |  |
| --- | --- | --- | --- | --- | --- | --- | --- | --- | --- |
|  | Unfavorable Discharge Disposition |  |  | Death at Discharge |  |  | Eye Tracking |  |  |
| Incidence Variables | OR | 95%CI | p | OR | 95%CI | P | OR | 95%CI | p |
| Pupil Abnormalities <sup>a</sup> | 1.62 | 0.51-5.7 | 0.424 | 4.78 | 0.96-38.34 | 0.085 | 0.16 | 0.02-0.82 | 0.05 |
| NPi < 3 | 1.75 | 0.66-4.67 | 0.26 | 2.45 | 0.8-7.91 | 0.122 | 0.41 | 0.13-1.25 | 0.121 |
| Diff NPi ≥ 0.7 | 1.1 | 0.4-2.93 | 0.85 | 1.06 | 0.33-3.46 | 0.917 | 0.79 | 0.25-2.5 | 0.687 |
| Diff Size ≥ 1 | 1.06 | 0.42-2.64 | 0.907 | 1.17 | 0.41-3.44 | 0.766 | 1.09 | 0.39-3.06 | 0.87 |
| Unilateral Pupil Abnormalities <sup>b</sup> | 1.17 | 0.4-3.58 | 0.782 | 1.56 | 0.42-6.99 | 0.526 | 0.5 | 0.11-1.89 | 0.334 |
| Bilateral Pupil Abnormalities <sup>c</sup> | 7.79 | 2.32-29.55 | 0.001 | 17.07 | 4.52-76.81 | <0.001 | 0.07 | 0.02-0.26 | <0.001 |

Multivariable logistic regression models were adjusted for age, race, severity group of traumatic brain injury, Marshall computed tomography score, and penetrating vs. blunt trauma. Incidence of bilateral pupil abnormality was associated with all clinical outcomes. While any pupil abnormality was not associated with unfavorable discharge disposition, it was associated with eye tracking.
**Abbreviations:** OR = Odds Ratio, CI = Confidence Interval, NPi = Average Neurological Pupil index (NPi) between both eyes, Diff NPi = Difference in NPi between both eyes, Diff Size = Difference in size between both eyes
<sup>a</sup>**Pupil Abnormalities:** NPi < 3, Diff NPi ≥ 0.7, or Diff Size ≥ 1mm
<sup>b</sup>**Unilateral Pupil Abnormalities:** Unilateral NPi < 3, Diff NPi ≥ 0.7, or Diff Size ≥ 1mm
<sup>c</sup>**Bilateral Pupil Abnormalities:** Bilateral NPi < 3

**eTable 6:** Multivariable Models of Pupil Abnormality Frequency within first 72 Hours and Discharge Outcomes.

|  | Primary Outcome |  |  | Secondary Outcome |  |  |  |  |  |
| --- | --- | --- | --- | --- | --- | --- | --- | --- | --- |
|  | Unfavorable Discharge Disposition |  |  | Death at Discharge |  |  | Eye Tracking |  |  |
| Frequency (median number of pupil measurements per patient) | OR | 95%CI | p | OR | 95%CI | p | OR | 95%CI | p |
| Pupil Abnormalities <sup>a</sup> | 1.03 | 1.01-1.05 | 0.002 | 1.05 | 1.03-1.07 | <0.001 | 0.96 | 0.93-0.98 | <0.001 |
| NPi < 3 | 1.05 | 1.02-1.1 | 0.012 | 1.07 | 1.04-1.12 | <0.001 | 0.94 | 0.9-0.97 | 0.001 |
| Diff NPi ≥ 0.7 | 1.01 | 0.99-1.03 | 0.349 | 1.01 | 0.99-1.04 | 0.278 | 0.99 | 0.96-1.01 | 0.227 |
| Diff Size ≥ 1 | 1.01 | 0.99-1.04 | 0.255 | 1.04 | 1.01-1.07 | 0.019 | 0.97 | 0.94-1 | 0.035 |
| Unilateral Pupil Abnormalities <sup>b</sup> | 1.02 | 1-1.04 | 0.132 | 1.02 | 1-1.05 | 0.047 | 0.98 | 0.96-1 | 0.044 |
| Bilateral Pupil Abnormalities <sup>c</sup> | 1.22 | 1.07-1.45 | 0.008 | 1.23 | 1.08-1.45 | 0.005 | 0.83 | 0.71-0.94 | 0.007 |

Multivariable logistic regression models were adjusted for age, race, severity group of traumatic brain injury, Marshall computed tomography score, and penetrating vs. blunt trauma. Pupil Abnormalities and bilateral pupil abnormalities are associated with unfavorable discharge disposition and death at discharge, while inversely associated with eye tracking.
**Abbreviations:** OR = Odds Ratio, CI = Confidence Interval, NPi = Average Neurological Pupil index (NPi) between both eyes, Diff NPi = Difference in NPi between both eyes, Diff Size = Difference in size between both eyes, Inf = Number > 10<sup>5</sup>
<sup>a</sup>**Pupil Abnormalities:** NPi < 3, Diff NPi ≥ 0.7, or Diff Size ≥ 1mm
<sup>b</sup>**Unilateral Pupil Abnormalities:** Unilateral NPi < 3, Diff NPi ≥ 0.7, or Diff Size ≥ 1mm
<sup>c</sup>**Bilateral Pupil Abnormalities:** Bilateral NPi < 3

**eTable 7:** Likelihood Ratio Tests of IMPACT Model with and without Abnormal Pupil Frequency.

| Primary Outcome Unfavorable Discharge Disposition |  |  |  |
| --- | --- | --- | --- |
| IMPACT Log Likelihood | IMPACT +<br>Abnormal Pupil Frequency<br>Log Likelihood | $\chi^2$ | *p* |
| - 48.46 | - 45.63 | 5.67 | 0.02 |
| Secondary Outcome Death at Discharge |  |  |  |
| IMPACT Log Likelihood | IMPACT +<br>Abnormal Pupil Frequency<br>Log Likelihood | $\chi^2$ | *p* |
| - 38.24 | - 30.75 | 14.98 | < 0.001 |

When predicting unfavorable discharge disposition and death at discharge, the IMPACT models with the addition of pupil abnormality frequency had improved goodness-of-fit compared to the IMPACT model alone.
**Abbreviations:** IMPACT = IMPACT TBI Model, Abnormal Pupil Frequency = Pupil Abnormalities Frequency

**eTable 8:**
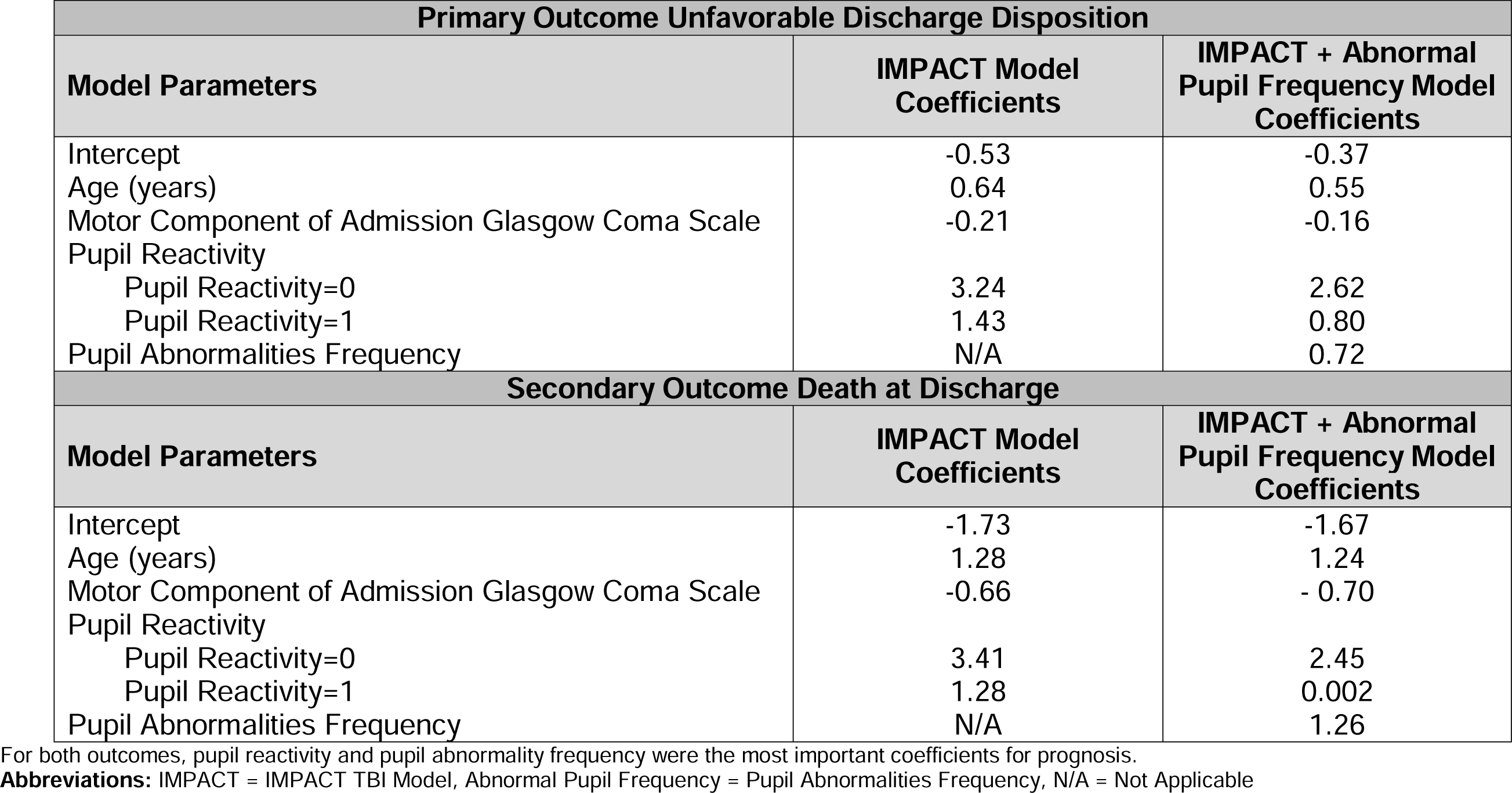
Parameters of IMPACT Models with and without Abnormal Pupil Frequency.

**eFigure 1:**
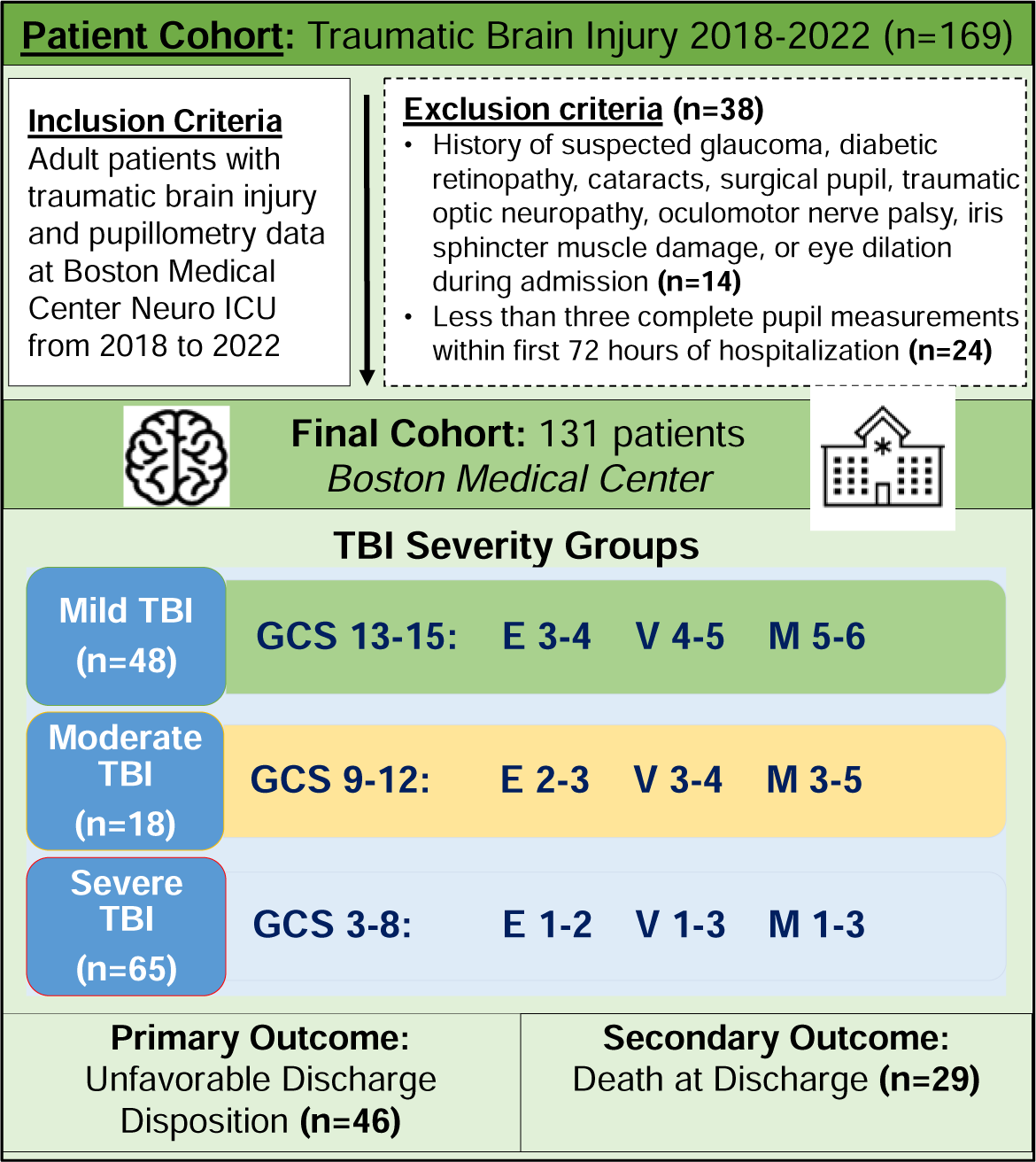
Inclusion and Exclusion Criteria. Inclusion and Exclusion Criteria for our study cohort of 131 patients. **Abbreviations:** ICU = Intensive Care Unit, TBI = Traumatic Brain Injury, GCS = Glasgow Coma Scale on Admission, E = Eye, V = Verbal, M = Movement **Unfavorable Discharge Disposition:** discharge to long-term care, hospice, or death

**eFigure 2:**
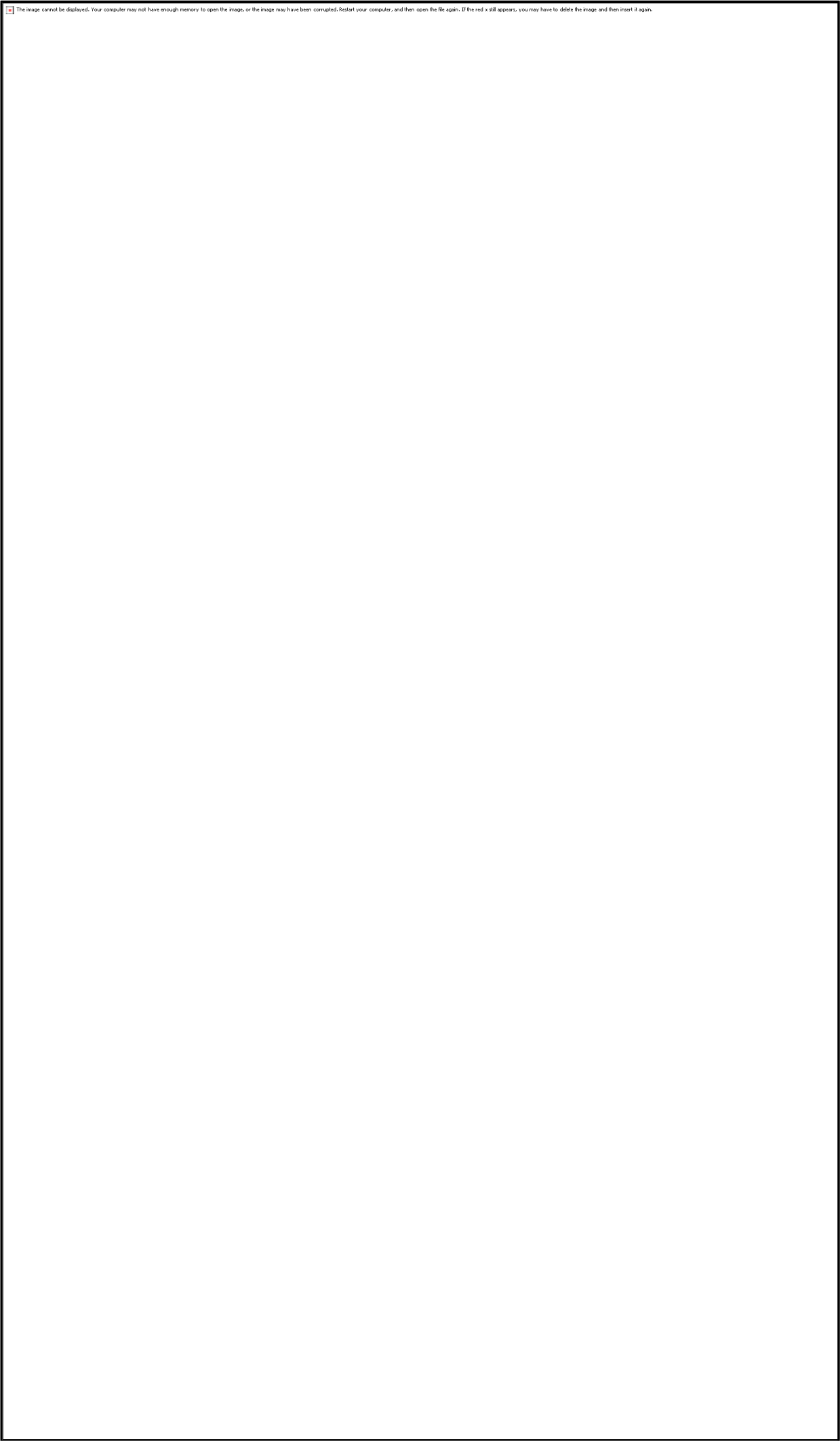
Violin Plots of Pupil Abnormality Frequency by Discharge Disposition. Distributions of pupil abnormalities for patients with at least one pupil abnormality. Pupil abnormality and bilateral pupil abnormality frequencies had higher medians and interquartile percentiles in unfavorable vs. favorable discharge disposition. For unilateral pupil abnormalities, patients with unfavorable discharge disposition had higher 75^th^ percentiles and more variable distribution compared to patients with favorable discharge disposition. **Abbreviations:** % = Percent

**eFigure 3:**
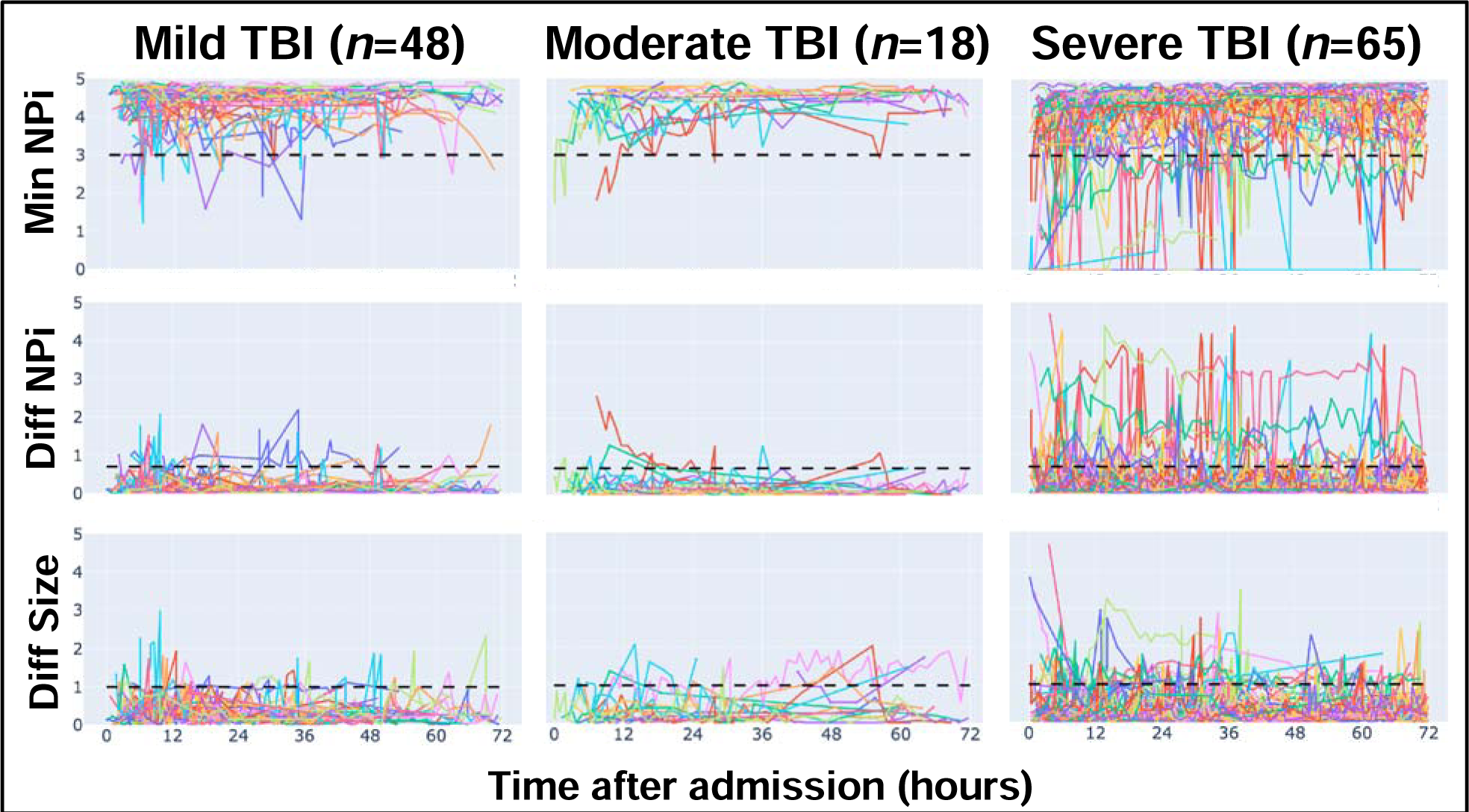
Minimum NPi, NPi Difference, and Size Difference by TBI Severity. Trajectories of minimum NPi, NPi difference, and size difference over the first 72 hours of hospitalization for each patient by traumatic brain injury (TBI) severity group. Each colored line represents a different patient. Thresholds for minimum NPi, NPi difference, and size difference are depicted in dashed black lines as 3, 0.7, and 1, respectively. There is an increased number of patients with abnormal minimum NPi, NPi difference, and size difference in patients with severe TBI compared to patients with mild or moderate TBI. **Abbreviations:** TBI = Traumatic Brain Injury, NPi = Neurological Pupil index, Min NPI = Minimum NPI between both eyes, Diff NPi = NPi difference between both eyes, Diff Size = Difference in resting pupil size between both eyes

**eFigure 4:**
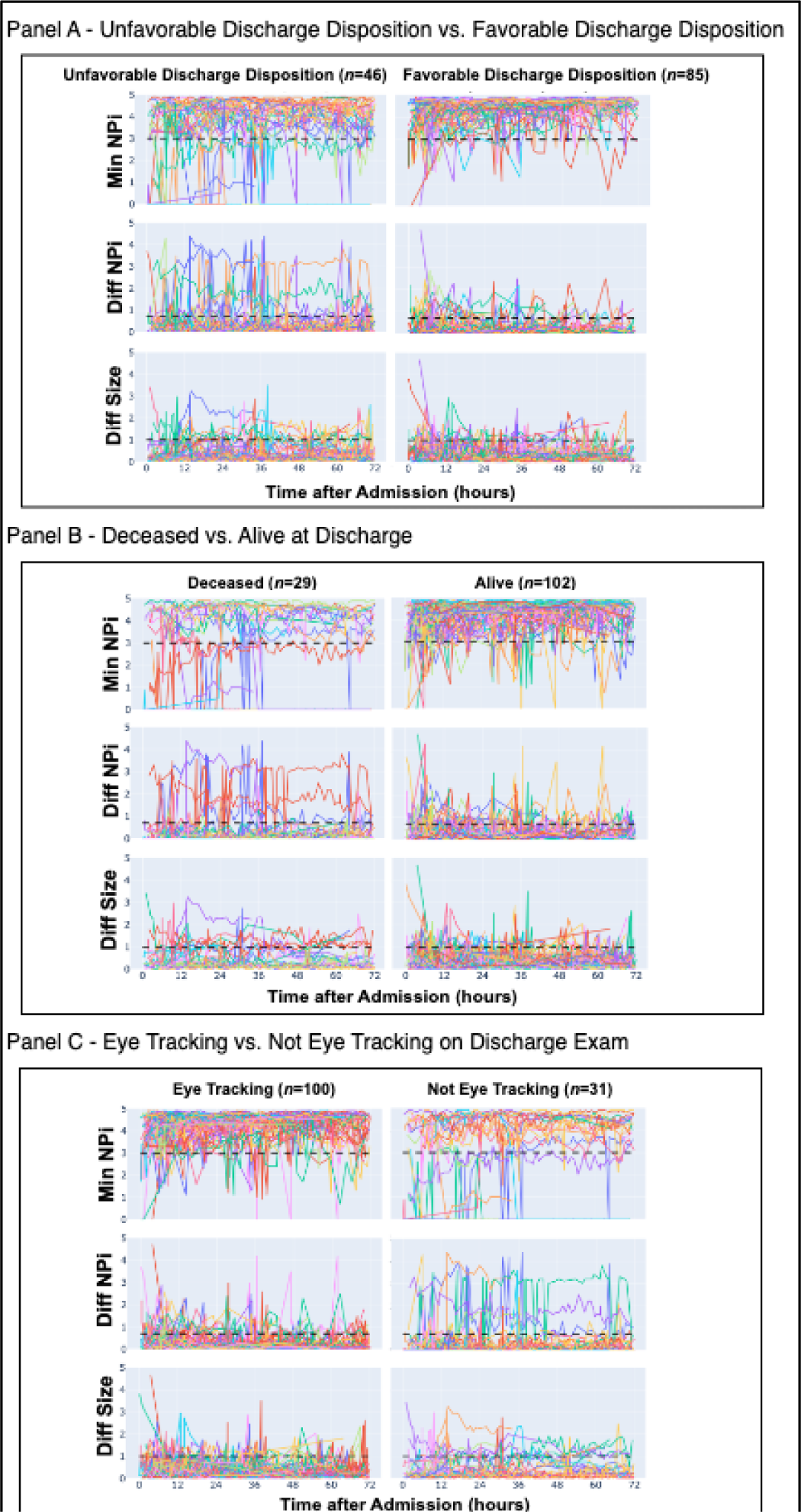
Minimum NPi, NPi Difference, and Size Difference by Discharge Outcomes. Trajectories of minimum NPi, NPi difference, and size difference for each patient over the first 72 hours of hospital stay by unfavorable vs. favorable discharge outcome, with thresholds for minimum NPi, NPi difference, and size difference depicted in dashed black lines as 3, 0.7, and 1, respectively. Unfavorable Discharge Disposition was defined as discharge disposition of deceased, hospice, or long-term care, while favorable discharge disposition was defined as discharge disposition of left against medical advice, home, skilled nursing facility, or rehab facility. **Abbreviations:** NPi = Neurological Pupil index, Min NPi = Minimum NPi between both eyes, Diff NPi = Difference in NPi between both eyes, Diff Size = Difference in resting pupil size between both eyes

**eFigure 5:**
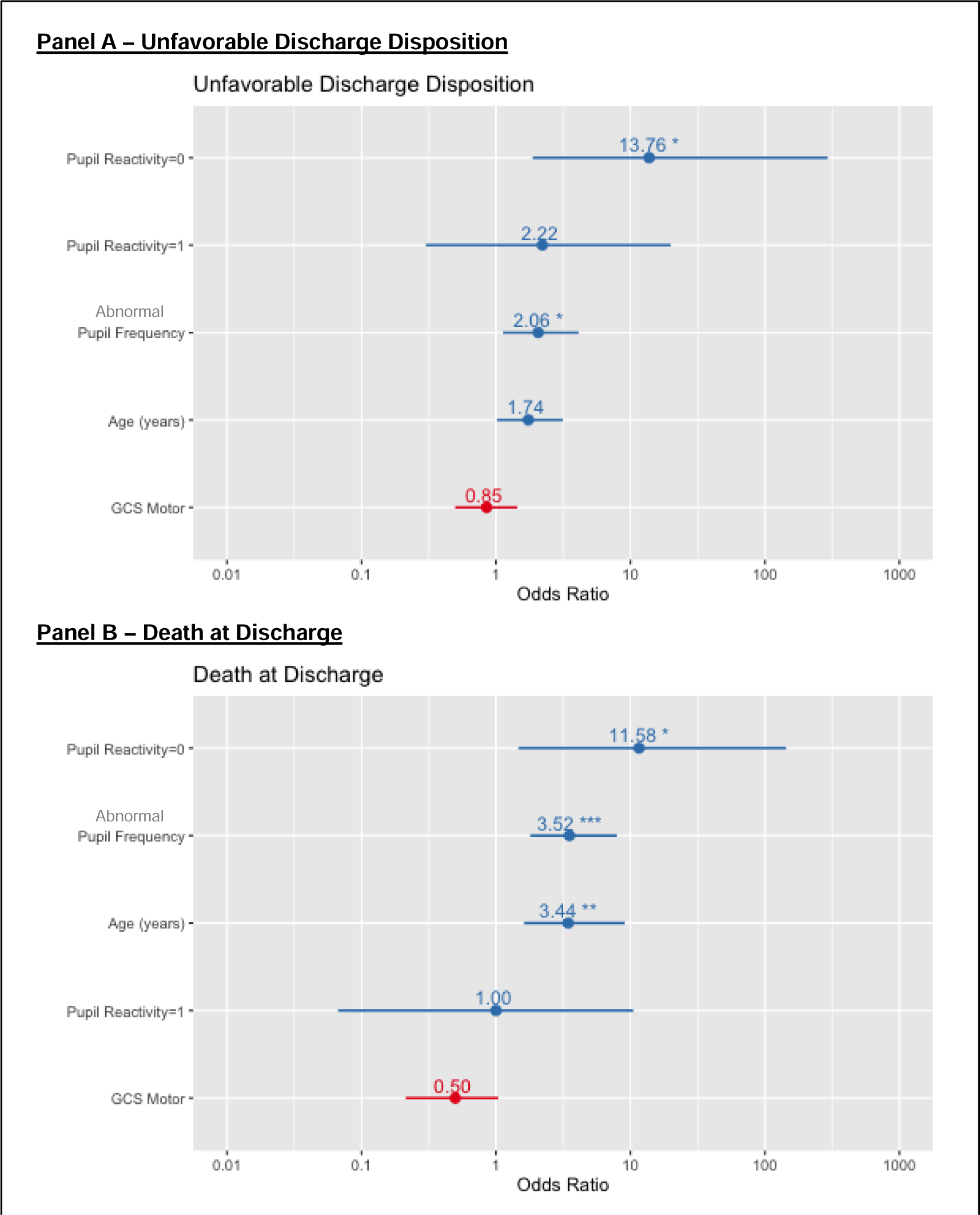
Odds Ratios for Parameters in IMPACT and Abnormal Pupil Frequency Models. Coefficients for features of the core IMPACT model trained using logistic regression analysis on our dataset. For both unfavorable discharge disposition and death at discharge, zero reactive pupils on admission and pupil abnormality frequency over the first 72 hours were the most important coefficients. **Abbreviations:** IMPACT = IMPACT TBI, Abnormal Pupil Frequency = Pupil Abnormalities Frequency, GCS = Glasgow Coma Scale

**eFigure 6:**
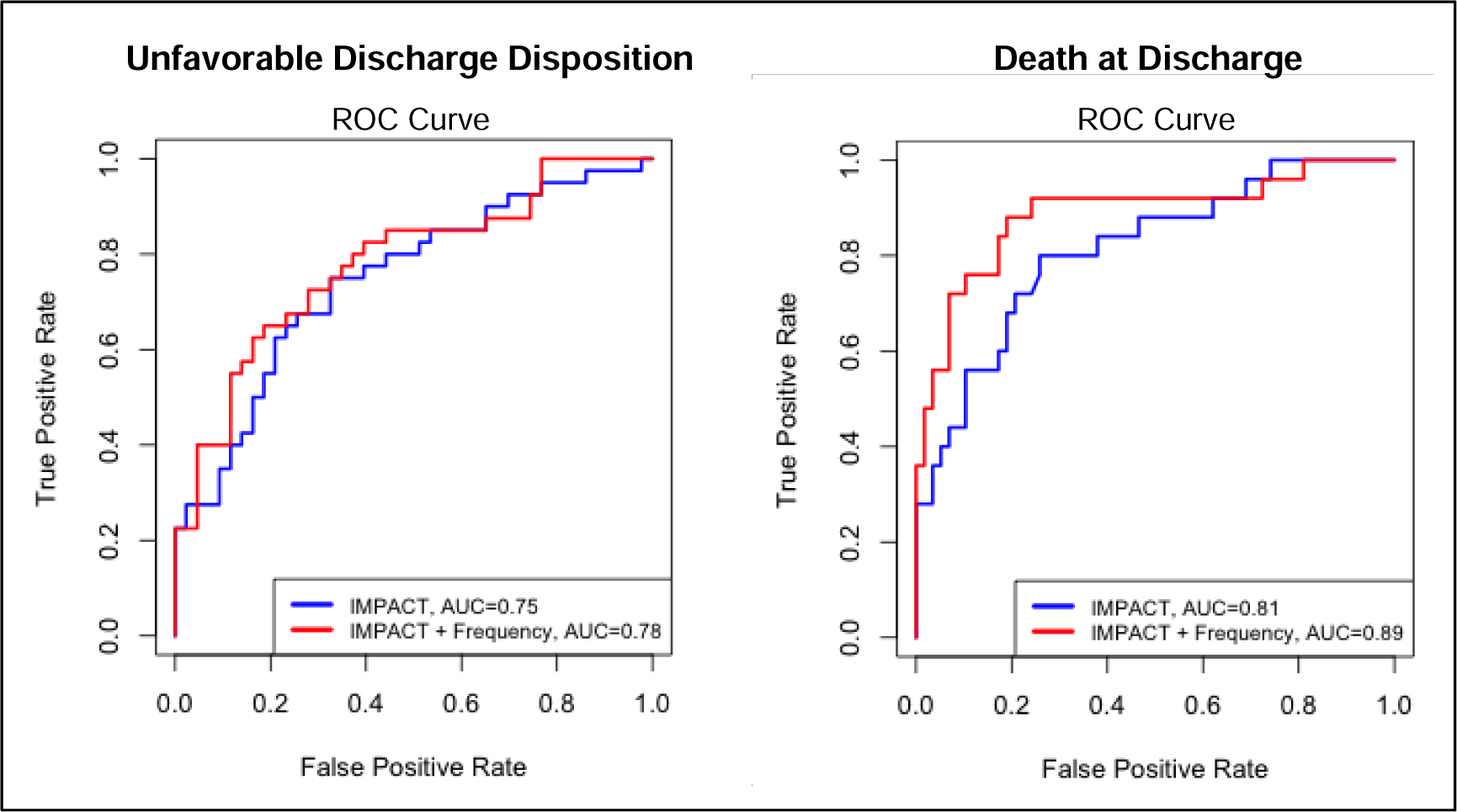
ROC Curves for IMPACT Model with and without Abnormal Pupil Frequency. ROC curves comparing IMPACT model with and without abnormal pupil frequency for both primary and secondary outcomes. For unfavorable discharge disposition, the IMPACT and abnormal pupil frequency model had improved area under the curve (AUC) (AUC = 0.78) compared to the IMPACT model alone (AUC = 0.75) although not statistically significant (DeLong’s test *p* = 0.2). For death at discharge, the IMPACT and abnormal pupil frequency model had improved area under the curve (AUC) (AUC = 0.89) compared to the IMPACT model alone (AUC = 0.81) although also not statistically significant (DeLong’s test *p* = 0.056). **Abbreviations:** ROC = receiver operating characteristic, IMPACT = IMPACT TBI, Frequency = Pupil Abnormalities Frequency, GCS = Glasgow Coma Scale

## References

1. Brazinova A, Rehorcikova V, Taylor MS, et al. Epidemiology of Traumatic Brain Injury in Europe: A Living Systematic Review. J Neurotrauma 2021;38(10):1411–1440, doi:10.1089/neu.2015.4126

2. El Ahmadieh TY, Bedros N, Stutzman SE, et al. Automated Pupillometry as a Triage and Assessment Tool in Patients with Traumatic Brain Injury. World Neurosurg 2021;145(e163-e169, doi:10.1016/j.wneu.2020.09.152

3. Hoffmann M, Lefering R, Rueger JM, et al. Pupil evaluation in addition to Glasgow Coma Scale components in prediction of traumatic brain injury and mortality. Br J Surg 2012;99 Suppl 1(122-30, doi:10.1002/bjs.7707

4. Marmarou A, Lu J, Butcher I, et al. Prognostic value of the Glasgow Coma Scale and pupil reactivity in traumatic brain injury assessed pre-hospital and on enrollment: an IMPACT analysis. J Neurotrauma 2007;24(2):270–80, doi:10.1089/neu.2006.0029

5. Han J, King NK, Neilson SJ, et al. External validation of the CRASH and IMPACT prognostic models in severe traumatic brain injury. J Neurotrauma 2014;31(13):1146–52, doi:10.1089/neu.2013.3003

6. Anderson M, Elmer J, Shutter L, et al. Integrating Quantitative Pupillometry Into Regular Care in a Neurotrauma Intensive Care Unit. J Neurosci Nurs 2018;50(1):30–36, doi:10.1097/JNN.0000000000000333

7. Alali AS, Temkin N, Barber J, et al. A clinical decision rule to predict intracranial hypertension in severe traumatic brain injury. J Neurosurg 2018;131(2):612–619, doi:10.3171/2018.4.JNS173166

8. Monai E, Favaretto C, Salvalaggio A, et al. Pupillary dynamics predict long-term outcome in a cohort of acute traumatic brain injury coma patients. Ann Clin Transl Neurol 2023;10(10):1854–1862, doi:10.1002/acn3.51879

9. Couret D, Boumaza D, Grisotto C, et al. Reliability of standard pupillometry practice in neurocritical care: an observational, double-blinded study. Crit Care 2016;20(99, doi:10.1186/s13054-016-1239-z

10. Oddo, M., Taccone, F. S., Petrosino, M., Badenes, R., Blandino-Ortiz, A., Bouzat, P., Caricato, A., Chesnut, R. M., Feyling, A. C., Ben-Hamouda, N., Hemphill, J. C., Koehn, J., Rasulo, F., Suarez, J. I., Elli, F., Vargiolu, A., Rebora, P., Galimberti, S., Citerio, G., & ORANGE study investigators (2023). The Neurological Pupil index for outcome prognostication in people with acute brain injury (ORANGE): a prospective, observational, multicentre cohort study. The Lancet. Neurology, 22(10), 925–933. 10.1016/S1474-4422(23)00271-5

11. Carney N, Totten AM, O’Reilly C, et al. Guidelines for the Management of Severe Traumatic Brain Injury, Fourth Edition. Neurosurgery 2017;80(1):6–15, doi:10.1227/NEU.0000000000001432

12. Marshall LF, Marshall SB, Klauber MR, et al. A new classification of head injury based on computerized tomography. Journal of Neurosurgery 1991;75(Supplement):S14–S20, 10.3171/sup.1991.75.1s.0s14

13. von Elm E, Altman DG, Egger M, et al. The Strengthening the Reporting of Observational Studies in Epidemiology (STROBE) statement: guidelines for reporting observational studies. Lancet 2007;370(9596):1453-7, doi:10.1016/S0140-6736(07)61602-X

14. Prescott BR, Saglam H, Duskin JA, et al. Anisocoria and Poor Pupil Reactivity by Quantitative Pupillometry in Patients With Intracranial Pathology. Crit Care Med 2022;50(2):e143–e153, doi:10.1097/CCM.0000000000005272

15. Amorim RL, Oliveira LM, Malbouisson LM, et al. Prediction of Early TBI Mortality Using a Machine Learning Approach in a LMIC Population. Front Neurol 2019;10(1366, doi:10.3389/fneur.2019.01366

16. Luz Teixeira T, Peluso L, Banco P, et al. Early Pupillometry Assessment in Traumatic Brain Injury Patients: A Retrospective Study. Brain Sci 2021;11(12), doi:10.3390/brainsci11121657

17. Turnbull BW, Weiss L. A likelihood ratio statistic for testing goodness of fit with randomly censored data. Biometrics 1978;34(3):367–75

18. Oddo M, Taccone FS, Petrosino M, et al. The Neurological Pupil index for outcome prognostication in people with acute brain injury (ORANGE): a prospective, observational, multicentre cohort study. Lancet Neurol 2023;22(10):925–933, doi:10.1016/S1474-4422(23)00271-5

19. Trent T, Vashisht A, Novakovic S, et al. Pupillary light reflex measured with quantitative pupillometry has low sensitivity and high specificity for predicting neuroworsening after traumatic brain injury. J Am Assoc Nurse Pract 2023;35(2):130–134, doi:10.1097/JXX.0000000000000822

20. Jahns FP, Miroz JP, Messerer M, et al. Quantitative pupillometry for the monitoring of intracranial hypertension in patients with severe traumatic brain injury. Crit Care 2019;23(1):155, doi:10.1186/s13054-019-2436-3

21. Romagnosi F, Bernini A, Bongiovanni F, et al. Neurological Pupil Index for the Early Prediction of Outcome in Severe Acute Brain Injury Patients. Brain Sci 2022;12(5), doi:10.3390/brainsci12050609

22. Larson MD, Behrends M. Portable infrared pupillometry: a review. Anesth Analg 2015;120(6):1242–53, doi:10.1213/ANE.0000000000000314

23. Diamond JP. The Pupil. Anatomy, Physiology and Clinical Applications. Br J Ophthalmol 2001;85(1):121E, doi:10.1136/bjo.85.1.121e

24. Steinhauer SR, Hakerem G. The pupillary response in cognitive psychophysiology and schizophrenia. Ann N Y Acad Sci 1992;658(182-204, doi:10.1111/j.1749-6632.1992.tb22845.x

## References

1. Harris PA, Taylor R, Thielke R, et al. Research electronic data capture (REDCap)--a metadata-driven methodology and workflow process for providing translational research informatics support. J Biomed Inform 2009;42(2):377–81, doi:10.1016/j.jbi.2008.08.010

2. Pollard TJ, Johnson AEW, Raffa JD, Mark RG. tableone: An open source Python package for producing summary statistics for research papers. JAMIA Open 2018;1(1):26–31, doi:10.1093/jamiaopen/ooy012

